# Genetically-proxied anti-diabetic drug target perturbation and risk of cancer: a Mendelian randomization analysis

**DOI:** 10.1101/2022.10.24.22281370

**Authors:** James Yarmolinsky, Emmanouil Bouras, Andrei Constantinescu, Kimberley Burrows, Caroline J Bull, Emma E Vincent, Richard M Martin, Olympia Dimopoulou, Sarah J Lewis, Victor Moreno, Marijana Vujkovic, Kyong-Mi Chang, Benjamin F Voight, Philip S Tsao, Marc J Gunter, Jochen Hampe, Annika Lindblom, Andrew J Pellatt, Paul D P Pharoah, Robert E Schoen, Steven Gallinger, Mark A Jenkins, Rish K Pai, the PRACTICAL consortium, VA Million Veteran Program, Dipender Gill, Kostas K Tsilidis

## Abstract

**Aims/hypothesis:** Epidemiological studies have generated conflicting findings on the relationship between anti-diabetic medication use and cancer risk. Naturally occurring variation in genes encoding anti-diabetic drug targets can be used to investigate the effect of their pharmacological perturbation on cancer risk.

**Methods:** We developed genetic instruments for three anti-diabetic drug targets (peroxisome proliferator activated receptor gamma, PPARG; sulfonylurea receptor 1, ABCC8; glucagon-like peptide 1 receptor, GLP1R) using summary genetic association data from a genome-wide association study (GWAS) of type 2 diabetes in 69,869 cases and 127,197 controls in the Million Veteran Program. Genetic instruments were constructed using *cis*-acting genome-wide significant (*P*<5×10^−8^) single-nucleotide polymorphisms (SNPs) permitted to be in weak linkage disequilibrium (r^2^<0.20). Summary genetic association estimates for these SNPs were obtained from GWAS consortia for the following cancers: breast (122,977 cases, 105,974 controls), colorectal (58,221 cases, 67,694 controls), prostate (79,148 cases, 61,106 controls), and overall (i.e. site-combined) cancer (27,483 cases, 372,016 controls). Inverse-variance weighted random-effects models adjusting for linkage disequilibrium were employed to estimate causal associations between genetically-proxied drug target perturbation and cancer risk. Colocalisation analysis was employed to examine robustness of findings to violations of Mendelian randomization (MR) assumptions. A Bonferroni correction was employed as a heuristic to define associations from MR analyses as “strong” and “weak” evidence.

**Results:** In Mendelian randomization analysis, genetically-proxied PPARG perturbation was weakly associated with higher risk of prostate cancer (OR for PPARG perturbation equivalent to a 1 unit decrease in inverse-rank normal transformed HbA_1c_: 1.75, 95% CI 1.07-2.85, *P*=0.02). In histological subtype-stratified analyses, genetically-proxied PPARG perturbation was weakly associated with lower risk of ER+ breast cancer (OR 0.57, 95% CI 0.38-0.85; *P*=6.45 × 10^−3^). In colocalisation analysis however, there was little evidence of shared causal variants for type 2 diabetes liability and cancer endpoints in the *PPARG* locus, though these analyses were likely underpowered. There was little evidence to support associations of genetically-proxied PPARG perturbation with colorectal or overall cancer risk or genetically-proxied ABCC8 or GLP1R perturbation with risk across cancer endpoints.

**Conclusions/interpretation:** Our drug-target MR analyses did not find consistent evidence to support an association of genetically-proxied PPARG, ABCC8 or GLP1R perturbation with breast, colorectal, prostate or overall cancer risk. Further evaluation of these drug targets using alternative molecular epidemiological approaches may help to further corroborate the findings presented in this analysis.

**Research in context:**

- What is already known about this subject?
  - Anti-diabetic medication use is variably linked to both increased and decreased cancer risk in conventional epidemiological studies
  - It is unclear whether these associations represent causal relationships
- What is the key question?
  - What is the association of genetically-proxied perturbation of three anti-diabetic drug targets (PPARG, ABCC8, GLP1R) with risk of breast, colorectal, prostate and overall cancer risk?
- What are the new findings?
  - Genetically-proxied PPARG perturbation was weakly associated with higher risk of prostate cancer and lower risk of ER+ breast cancer
  - There was little evidence that liability to type 2 diabetes and these cancer endpoints shared one or more causal variants in the *PPARG* locus, a necessary precondition to infer causality between PPARG perturbation and cancer risk
- How might this impact on clinical practice in the foreseeable future?
  - Our drug-target Mendelian randomization analyses did not find consistent evidence to support a link between genetically-proxied perturbation of PPARG, ABCC8, and GLP1R and risk of breast, colorectal, prostate and overall cancer risk
  - These findings suggest that on-target effects of PPARG agonists, sulfonylureas, and GLP1R agonists are unlikely to confer large effects on breast, colorectal, prostate, or overall cancer risk

## Introduction

Globally, an estimated 460 million individuals have type 2 diabetes, the majority of whom require long-term use of anti-diabetic medications to maintain glycaemic control [1]. Several different classes of oral anti-diabetic medications are used to manage this condition, including biguanides (e.g. metformin), sulphonylureas, thiazolidinediones, dipeptidyl peptidase-4 (DPP-4) inhibitors, sodium-glucose cotransporter 2 (SGLT2) inhibitors and glucagon-like peptide-1 receptor (GLP1R) agonists, with diverse mechanisms of action [2].

Preclinical studies have variably reported both carcinogenic and anti-neoplastic effects of anti-diabetic medications. For example, *in vitro* studies have suggested that metformin, an insulin sensitiser and first-line therapy for type 2 diabetes, can reduce cell proliferation, induce apoptosis, and cause cell cycle arrest [3]. Thiazolidinediones, insulin sensitisers and selective peroxisome proliferator-activator nuclear receptor (PPARG) agonists, have been suggested to increase cellular differentiation, reduce cellular proliferation, and induce apoptosis in some cell lines but to promote metastatic prostate cancer *in vivo* [4-6]. There is also some evidence that sulphonylureas, secretagogues that lower blood glucose levels by stimulating pancreatic insulin secretion, may promote carcinogenesis, potentially via increasing circulating insulin levels [7, 8]. Finally, *in vitro* studies have reported potential antiproliferative effects of GLP1R agonists in various cancer cell types[9-11].

Epidemiological studies of anti-diabetic medication use have provided some support for findings from laboratory studies. For example, some observational studies have reported that metformin users have lower risk of several cancers while sulphonylurea use has been associated with an increased risk of site-specific (i.e. colorectal, metastatic prostate) and overall cancer [12-17]. In addition, some thiazolidinediones (i.e. pioglitazones) have been linked to an elevated risk of bladder, prostate, and pancreatic cancer, though use of rosiglitazones has been associated with lower breast cancer risk [18, 19]. Finally, GLP1R agonist use has been associated with a decreased risk of prostate cancer when compared with sulfonylurea use [20].

The causal nature of associations reported between anti-diabetic medication use and cancer risk in conventional epidemiological studies is often unclear. This is because of the susceptibility of such studies to residual confounding (e.g. due to indication) and various forms of bias (e.g. immortal time, prevalent user), which can undermine robust causal inference [21-23]. While clinical trials of anti-diabetic medications have not consistently reported differences in rates of cancer among users of these medications, such studies are often underpowered to detect effects for individual cancer sites [23-27]. Further, such studies often have limited follow-up periods, thus not being able to adequately capture outcomes with long induction periods, such as cancer.

Drug target-Mendelian randomization (MR) uses germline variants in genes encoding drug targets as instruments (“proxies”) for these targets to estimate the effect of their pharmacological perturbation on disease endpoints [28, 29]. Since germline genetic variants are randomly assorted at meiosis and fixed at conception, analyses using variants as instruments should be less prone to conventional issues of confounding and reverse causation. In addition, given the length of time required for solid tumour development, the use of germline genetic variants as instruments is advantageous as it permits estimation of the long-term effects of medication use on cancer risk [30].

Given the widespread use of anti-diabetic medications and reports of both adverse and protective associations of these medications with cancer risk in preclinical and epidemiological studies, there is a need to further evaluate the role of these medications in the risk of common adult cancers. Additionally, given the long induction periods of cancer, using MR to examine target-mediated effects of medications that have been on the market for relatively short periods of time (e.g. SGLT2 inhibitors and GLP1R agonists) can be informative in predicting their long-term safety profiles. We thus aimed to i) develop genetic instruments for the targets of five approved type 2 diabetes medications with known mechanisms of action [sulfonylurea receptor 1 (ABCC8), PPARG, sodium/glucose cotransporter 2 (SLC5A2), DPP4, GLP1R)] and ii) evaluate associations of genetically-proxied perturbation of three of these targets with reliable *cis*-acting instruments (ABCC8, PPARG, and GLP1R) with risk of breast, colorectal, prostate, common cancers with epidemiological evidence suggesting a link between anti-diabetic medication use and their onset, and overall (i.e. site-combined) cancer [5, 12-14, 18, 19, 31-33].

## Methods

Summary genetic association data were obtained from three cancer-specific genome-wide association study (GWAS) consortia. Summary genetic association estimates for overall and oestrogen receptor-stratified breast cancer risk in up to 122,977 cases and 105,974 controls were obtained from the Breast Cancer Association Consortium (BCAC) [34]. Summary genetic association estimates for overall and site-specific (i.e. colon, rectal) colorectal cancer risk in up to 58,221 cases and 67,694 controls were obtained from an analysis of the Genetics and Epidemiology of Colorectal Cancer Consortium (GECCO), ColoRectal Transdisciplinary Study (CORECT), and Colon Cancer Family Registry (CCFR) [35]. Summary genetic association estimates for overall and advanced prostate cancer risk (i.e. metastatic disease, Gleason score ≥8, prostate-specific antigen >100 or prostate cancer-related death) in up to 79,148 cases and 61,106 controls were obtained from the Prostate Cancer Association Group to Investigate Cancer Associated Alterations in the Genome (PRACTICAL) consortium [36]. These analyses were restricted to participants of European ancestry.

Overall (i.e. site-combined) cancer risk data in 27,483 incident cases and 372,016 controls were also obtained from a GWAS performed in the UK Biobank cohort study [37]. Briefly, cancer cases were classified according to the International Classification of Diseases (ICD9, ICD10) with data completed to April 2019 and controls were defined as individuals who did not have any cancer code (ICD9 or ICD10) and did not self-report a cancer diagnosis. GWAS were performed using a linear mixed model as implemented in BOLT-LMM (v2.3) (to account for relatedness and population stratification) and adjusted for age, sex, and genotyping array [38, 39].

Further information on statistical analysis, imputation, and quality control measures for these studies is available in the original publications. All studies contributing data to these analyses had the relevant institutional review board approval from each country, in accordance with the Declaration of Helsinki, and all participants provided informed consent.

### Instrument construction

To generate genetic instruments to proxy anti-diabetic drug target perturbation, summary genetic association data were obtained from a GWAS of type 2 diabetes (T2D) in the Million Veteran Program (148,726 cases; 965,732 controls of European ancestry) [40]. Analyses were adjusted for age, sex, and ten principal components of genetic ancestry. Instruments were constructed in PLINK by obtaining SNPs associated with type 2 diabetes at genome-wide significance (*P* < 5 × 10^−8^) that were in or within ±500 kb from the gene encoding each respective target (PPARG, Chr3: 12328867-12475855; ABCC8, Chr11: 17414432-17498449; GLP1R, Chr6: 39016574-39055519) using the 1000 Genomes Phase 3 reference panel [41, 42]. We were unable to identify genome-wide significant SNPs within 500kb windows from *SLC5A2* and *DPP4* (i.e. instruments for SGLT2 and DPP-4 inhibitors, respectively) and therefore did not proceed with Mendelian randomization analyses for these targets. We also did not include metformin as a drug target due to the unclear mechanism(s) of action of this medication [43]. For PPARG, ABCC8, and GLP1R, SNPs used as instruments were permitted to be in weak linkage disequilibrium (r^2^<0.20) with each other to increase the proportion of variance in each respective drug target explained by the instrument, maximising instrument strength [44]. In total, 9 SNPs that met these criteria were obtained for PPARG, 6 for ABCC8, and 4 for GLP1R.

In a separate population, we then evaluated the association of type 2 diabetes SNPs in drug target regions with glycated haemoglobin (HbA_1c_) levels, a marker of long-term blood glucose levels, in order to minimise winner’s curse bias. SNP summary statistics were re-scaled to represent a mmol/mol unit reduction in HbA_1c_ to provide more interpretable effect estimates in Mendelian randomization analyses. HbA_1c_ values were obtained from a GWAS of 407,766 participants of the UK Biobank study and adjusted for age, sex, batch, and ten principal components of genetic ancestry.

For the PPARG instrument, 2 SNPs where the effect on HbA_1c_ was in the opposite direction to that of type 2 diabetes were removed from the instrument (rs17036160, rs11712085), as these associations likely represent pleiotropic mechanisms that would bias consequent Mendelian randomization analyses.

### Instrument validation

Instruments were validated by examining the association of genetically-proxied drug target perturbation with endpoints influenced by these medications in randomised controlled trials. For PPARG, alanine aminotransferase (ALT) and aspartate aminotransferase (AST) levels were used as positive controls and for ABCC8 and GLP1R, body mass index (BMI) was used [45-48]. Colocalisation was then performed to assess whether drug targets and traits representing positive controls share the same causal variant at a given locus. Such an analysis can permit exploration of whether drug targets and positive control traits are influenced by distinct causal variants that are in linkage disequilibrium with each other, indicative of horizontal pleiotropy (an instrument influencing an outcome through pathways independent to that of the exposure), a violation of the exclusion restriction criterion [49].

Colocalisation analysis was performed using the coloc R package which uses approximate Bayes factor computation to generate posterior probabilities that associations between two traits represent each of the following configurations: (i) neither trait has a genetic association in the region (H_0_), (ii) only the first trait has a genetic association in the region (H_1_), (iii) only the second trait has a genetic association in the region (H_2_), (iv) both traits are associated but have different causal variants (H_3_) and (v) both traits are associated and share a single causal variant (H_4_) [50]. Colocalisation analysis was performed by generating ±500□Dkb windows around the gene encoding each respective drug target. We used a posterior probability of >0.50 to indicate support for a configuration tested [51]. Where there was not support for H_4_, we then examined the possibility of colocalisation across other secondary conditionally independent signals for either drug targets or positive controls by performing pairwise conditional and colocalization analysis on all conditionally independent association signals using GCTA-COJO and the coloc package as implemented in pwCoCo[50, 52, 53].

### Statistical analysis

Causal estimates were generated using inverse-variance weighted (IVW) random-effects models (permitting overdispersion in models). These models were adjusted for weak linkage disequilibrium between SNPs with reference to the 1000 Genomes Phase 3 reference panel [41, 54]. Where there was under-dispersion in causal estimates generated from individual genetic variants, the residual standard error was set to 1 (i.e. equivalent to a fixed-effects model).

Mendelian randomization analysis assumes that a genetic instrument (i) is associated with a modifiable exposure or drug target (“relevance”), (ii) does not share a common cause with an outcome (“exchangeability”), and (iii) has no direct effect on the outcome (“exclusion restriction”)[28].

The “relevance” MR assumption was evaluated by generating estimates of the proportion of variance of each drug target (in HbA_1c_ units) explained by the instrument (r^2^) and F-statistics. F-statistics can be used to examine whether results are likely to be influenced by weak instrument bias, i.e., reduced statistical power and bias when an instrument explains a limited proportion of the variance in a drug target. As a convention, a F-statistic of >10 is indicative of minimal weak instrument bias [55].

We evaluated the “exclusion restriction” Mendelian randomization assumption by performing colocalisation to examine whether drug targets and cancer endpoints showing nominal evidence of an association in MR analyses (*P* < 0.05) share the same causal variant at a given locus. Iterative leave-one-out analysis was performed iteratively removing one SNP at a time from instruments to examine whether findings showing nominal evidence of association were driven by a single influential SNP.

To account for multiple testing across analyses, a Bonferroni correction was used to establish a *P*-value threshold of < 0.0019 (false positive rate = 0.05/27 statistical tests [3 drug targets tested against 9 primary cancer endpoints]), which we used as a heuristic to define “strong evidence”, with findings between *P* ≥ 0.0019 and *P* < 0.05 defined as “weak evidence”.

## Results

Characteristics of genetic variants used to instrument anti-diabetic drug targets are presented in **Table 1**. Across all three drug targets, F-statistics for their respective instruments ranged from 34.4 to 583.0, suggesting that weak instrument bias was unlikely to affect the conclusions (**Supplementary Table 1**). Power calculations suggested that we had 80% power to detect ORs ranging from 1.40-2.62 (in PPARG analyses), 2.03-8.34 (in ABCC8 analyses), and 2.22-8.78 (in GLP1R analyses) per mmol/mol reduction in target-mediated inverse rank-normal transformed HbA_1c_ across all cancer endpoints (α=0.05). Complete power estimates across all MR analyses are presented in **Supplementary Table 2**.

**Table 1.**
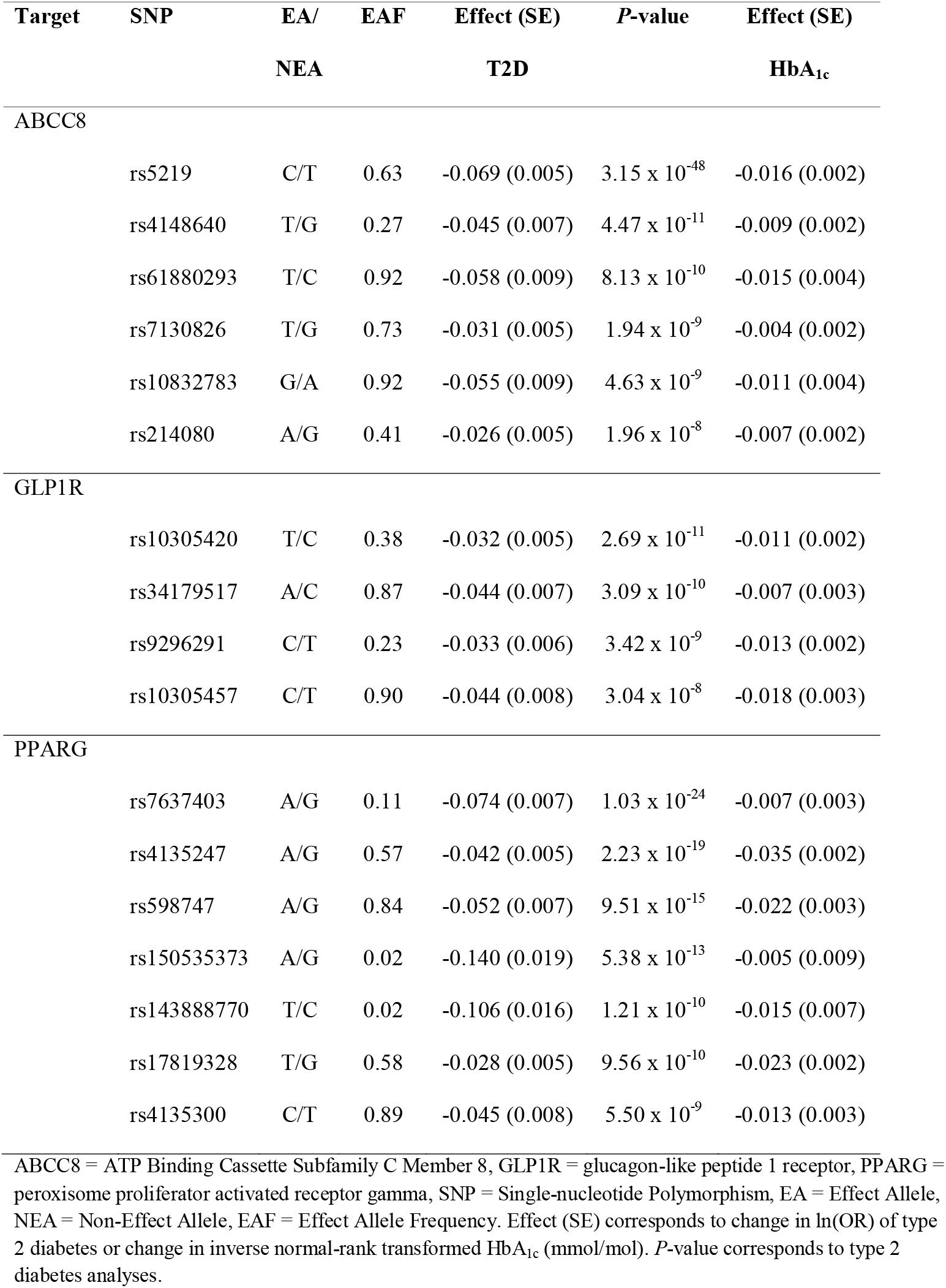
Characteristics of single-nucleotide polymorphisms used as instruments to proxy drug targets

| Target | SNP | EA/<br>NEA | EAF | Effect (SE) | P-value | Effect (SE) |
| --- | --- | --- | --- | --- | --- | --- |
|  |  |  |  | T2D |  | HbA<sub>1c</sub> |
| ABCC8 |  |  |  |  |  |  |
|  | rs5219 | C/T | 0.63 | -0.069 (0.005) | 3.15 x 10<sup>-48</sup> | -0.016 (0.002) |
|  | rs4148640 | T/G | 0.27 | -0.045 (0.007) | 4.47 x 10<sup>-11</sup> | -0.009 (0.002) |
|  | rs61880293 | T/C | 0.92 | -0.058 (0.009) | 8.13 x 10<sup>-10</sup> | -0.015 (0.004) |
|  | rs7130826 | T/G | 0.73 | -0.031 (0.005) | 1.94 x 10<sup>-9</sup> | -0.004 (0.002) |
|  | rs10832783 | G/A | 0.92 | -0.055 (0.009) | 4.63 x 10<sup>-9</sup> | -0.011 (0.004) |
|  | rs214080 | A/G | 0.41 | -0.026 (0.005) | 1.96 x 10<sup>-8</sup> | -0.007 (0.002) |
| GLP1R |  |  |  |  |  |  |
|  | rs10305420 | T/C | 0.38 | -0.032 (0.005) | 2.69 x 10<sup>-11</sup> | -0.011 (0.002) |
|  | rs34179517 | A/C | 0.87 | -0.044 (0.007) | 3.09 x 10<sup>-10</sup> | -0.007 (0.003) |
|  | rs9296291 | C/T | 0.23 | -0.033 (0.006) | 3.42 x 10<sup>-9</sup> | -0.013 (0.002) |
|  | rs10305457 | C/T | 0.90 | -0.044 (0.008) | 3.04 x 10<sup>-8</sup> | -0.018 (0.003) |
| PPARG |  |  |  |  |  |  |
|  | rs7637403 | A/G | 0.11 | -0.074 (0.007) | 1.03 x 10<sup>-24</sup> | -0.007 (0.003) |
|  | rs4135247 | A/G | 0.57 | -0.042 (0.005) | 2.23 x 10<sup>-19</sup> | -0.035 (0.002) |
|  | rs598747 | A/G | 0.84 | -0.052 (0.007) | 9.51 x 10<sup>-15</sup> | -0.022 (0.003) |
|  | rs150535373 | A/G | 0.02 | -0.140 (0.019) | 5.38 x 10<sup>-13</sup> | -0.005 (0.009) |
|  | rs143888770 | T/C | 0.02 | -0.106 (0.016) | 1.21 x 10<sup>-10</sup> | -0.015 (0.007) |
|  | rs17819328 | T/G | 0.58 | -0.028 (0.005) | 9.56 x 10<sup>-10</sup> | -0.023 (0.002) |
|  | rs4135300 | C/T | 0.89 | -0.045 (0.008) | 5.50 x 10<sup>-9</sup> | -0.013 (0.003) |

ABCC8 = ATP Binding Cassette Subfamily C Member 8, GLP1R = glucagon-like peptide 1 receptor, PPARG = peroxisome proliferator activated receptor gamma, SNP = Single-nucleotide Polymorphism, EA = Effect Allele, NEA = Non-Effect Allele, EAF = Effect Allele Frequency. Effect (SE) corresponds to change in ln(OR) of type 2 diabetes or change in inverse normal-rank transformed HbA<sub>1c</sub> (mmol/mol). P-value corresponds to type 2 diabetes analyses.

### Instrument validation

Genetically-proxied PPARG perturbation was associated with lower levels of ALT (SD change in ALT per PPARG perturbation equivalent to 1 unit inverse-rank normal transformed [IRNT] HbA_1c_ reduction: -0.57, 95% CI -1.01, -0.13; *P*=0.01) and AST (−0.49, 95% CI -1.79, -0.19; *P*=1.53 × 10^−3^). Colocalisation analysis suggested that T2D associations in the *PPARG* locus had a 91.8% and 83.9% probability of sharing a causal variant with ALT and AST, respectively (**Supplementary Tables 3-4, Figures 1-3**).

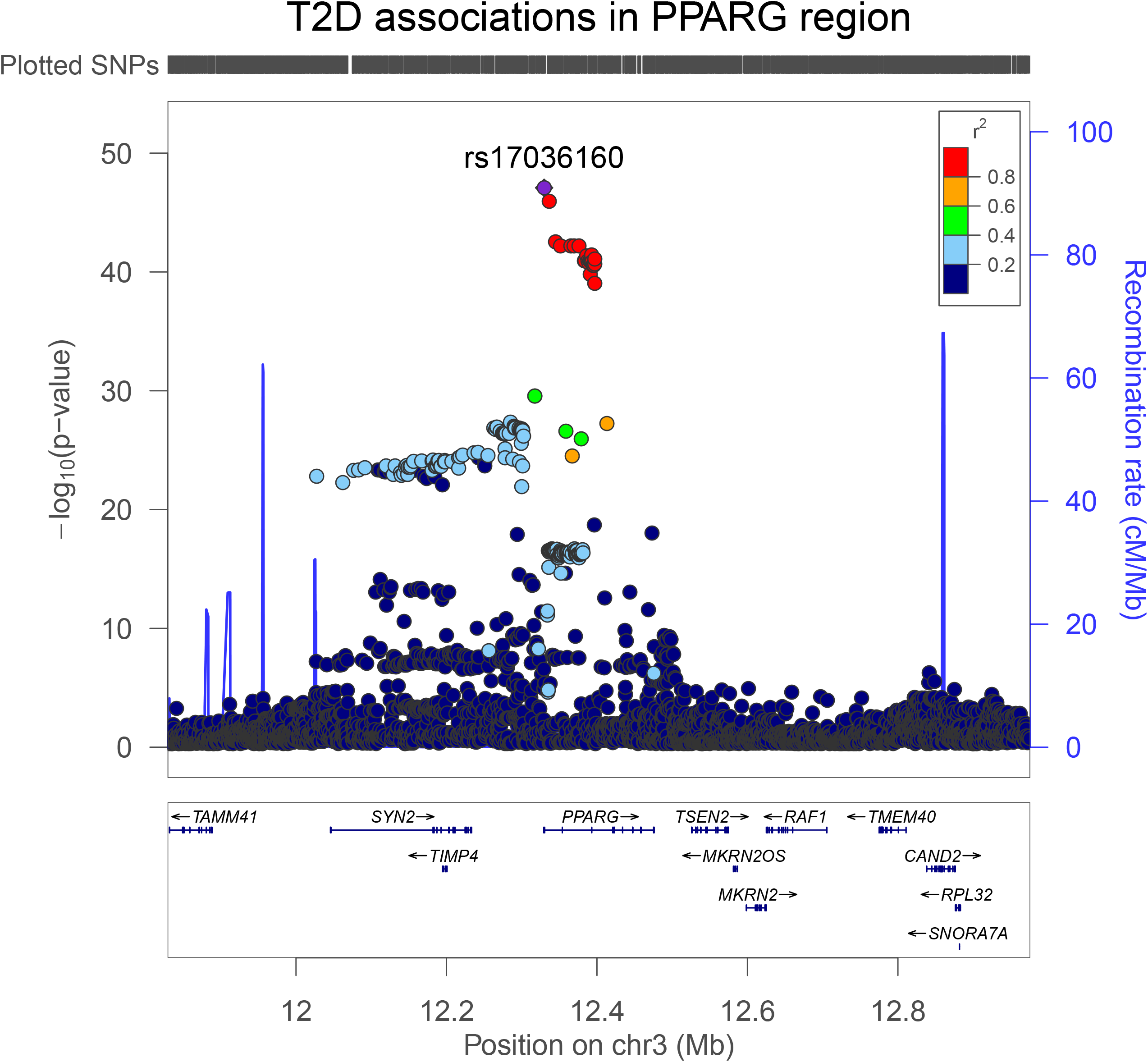

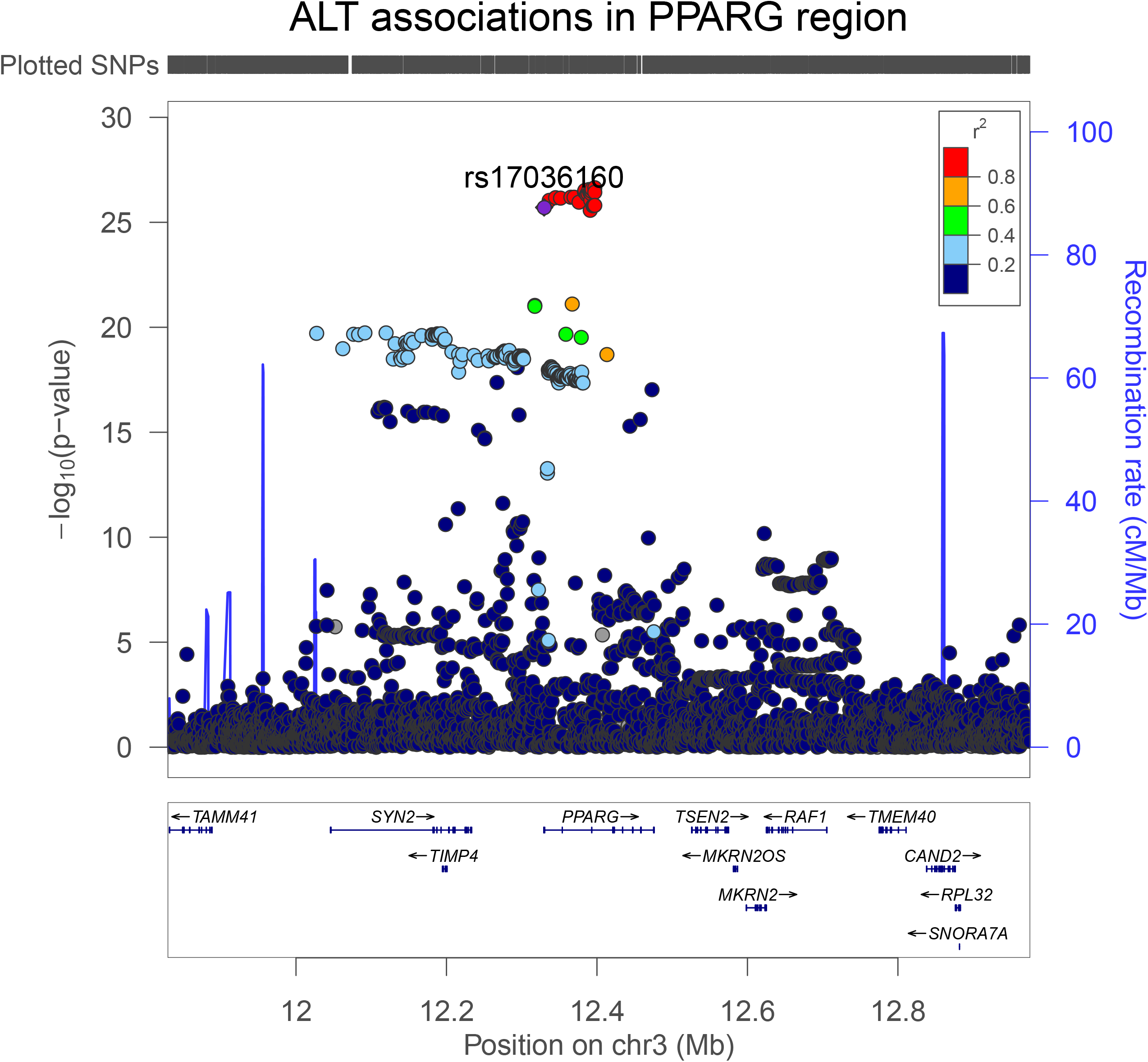

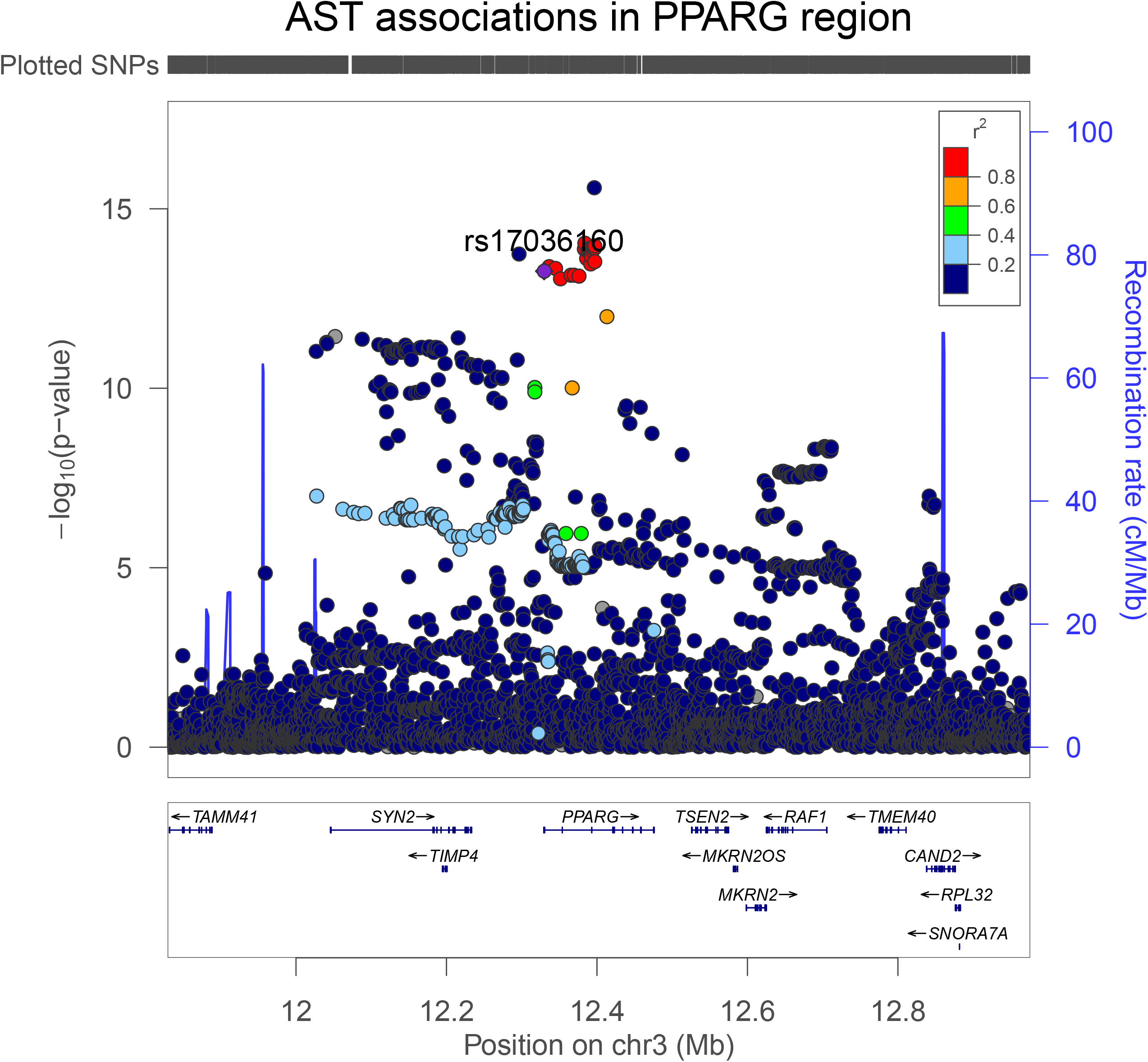

Genetically-proxied ABCC8 perturbation was associated with elevated BMI (SD change in BMI per ABCC8 perturbation equivalent to 1 unit IRNT HbA_1c_ reduction: 0.530, 95% CI 0.004, 0.172; *P*=3.75 × 10^−3^). Colocalisation analysis suggested that ABCC8 and BMI had a 94.0% posterior probability of sharing a causal variant within the *ABCC8* locus (**Supplementary Table 5, Figures 4-5**).

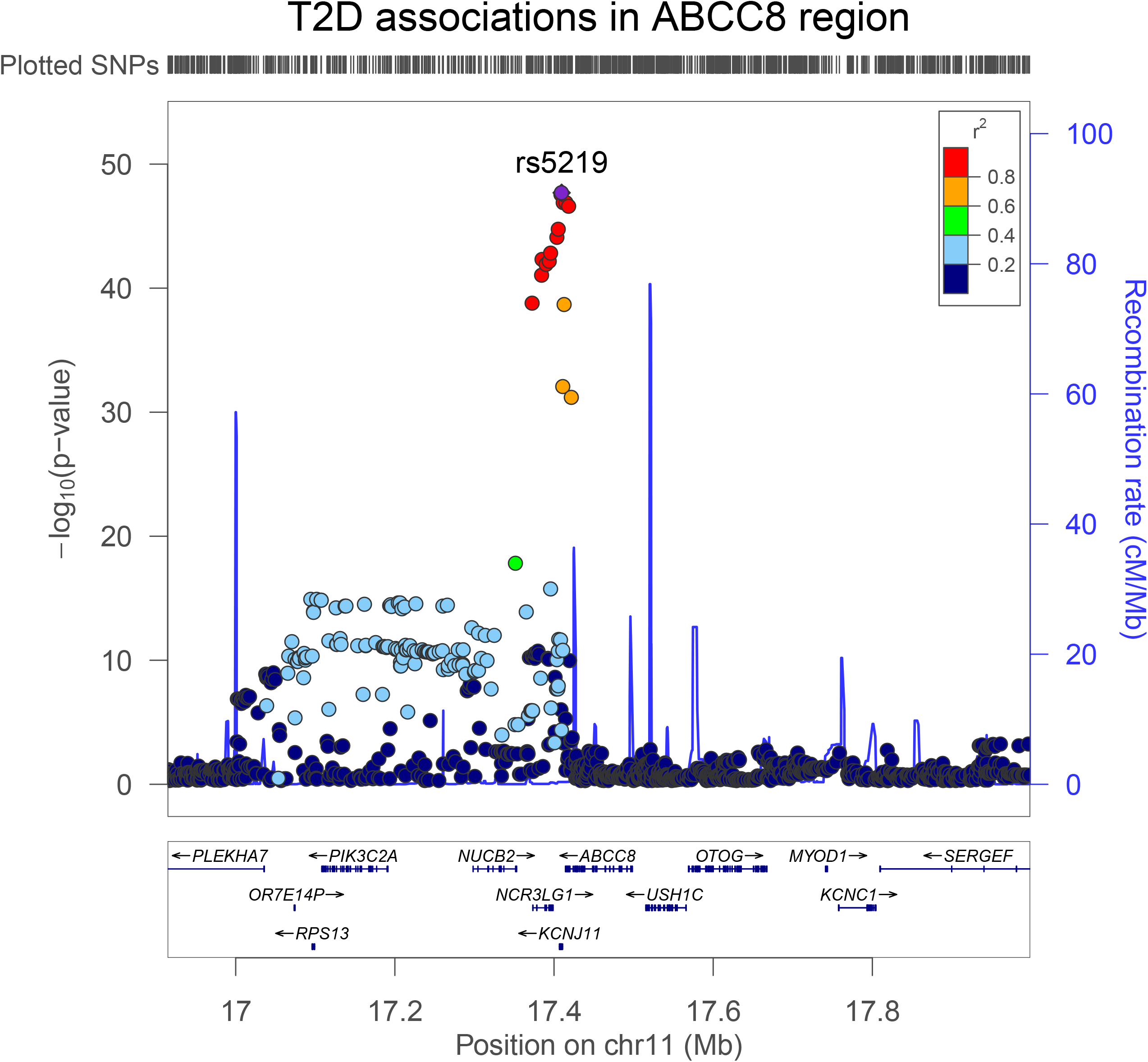

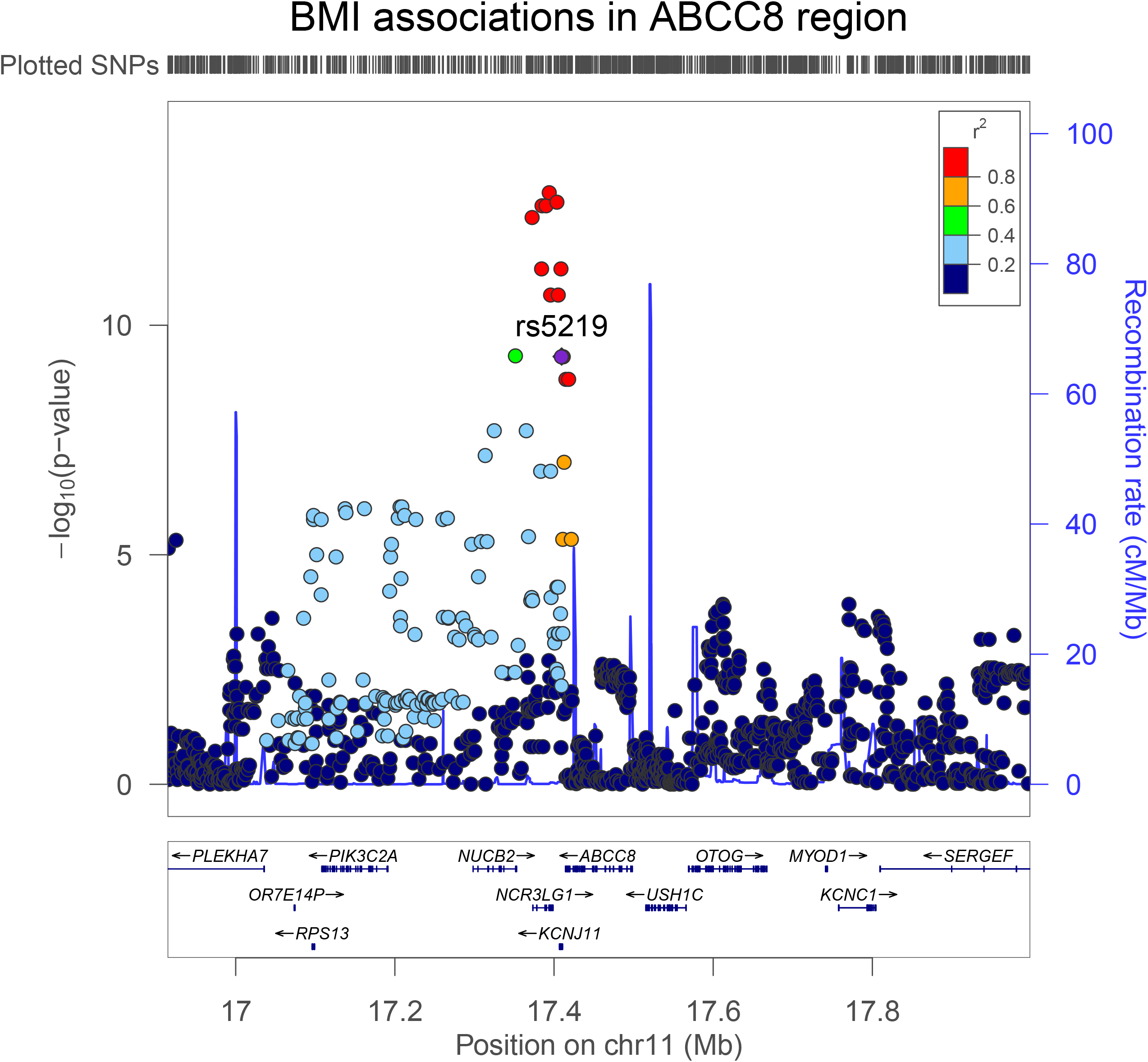

There was little evidence to support an association of genetically-proxied GLP1R perturbation with BMI (SD change in BMI equivalent to 1 unit IRNT HbA_1c_ reduction: -0.08, 95% CI -0.30, 0.15; *P*=0.51). Colocalisation analysis applied to both marginal and conditionally independent associations for T2D and BMI in the *GLP1R* locus did not support shared causal variants across these traits (posterior probability of shared causal variants across models: 0.22-0.49%) (**Supplementary Table 6, Figures 6-7**).

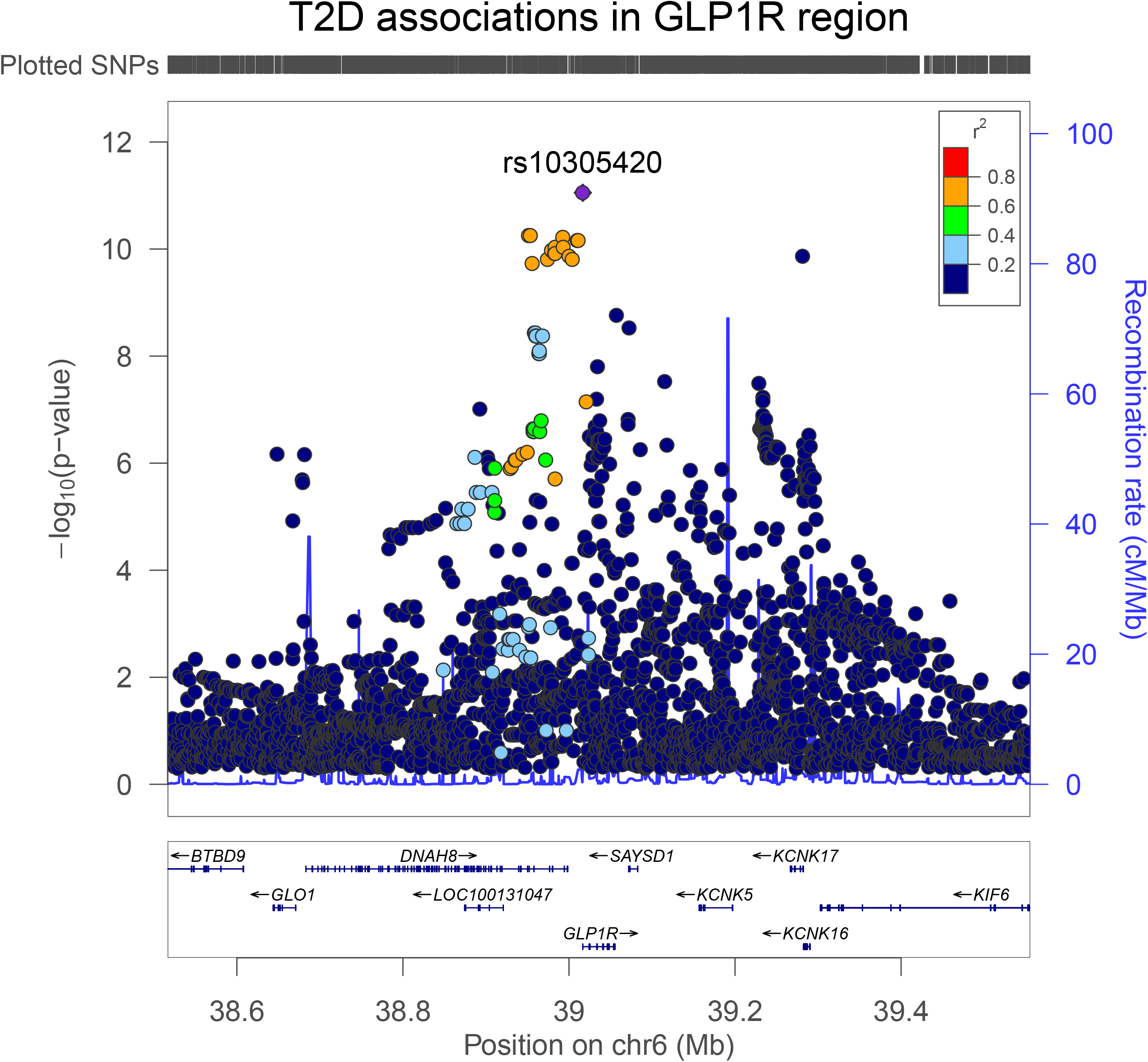

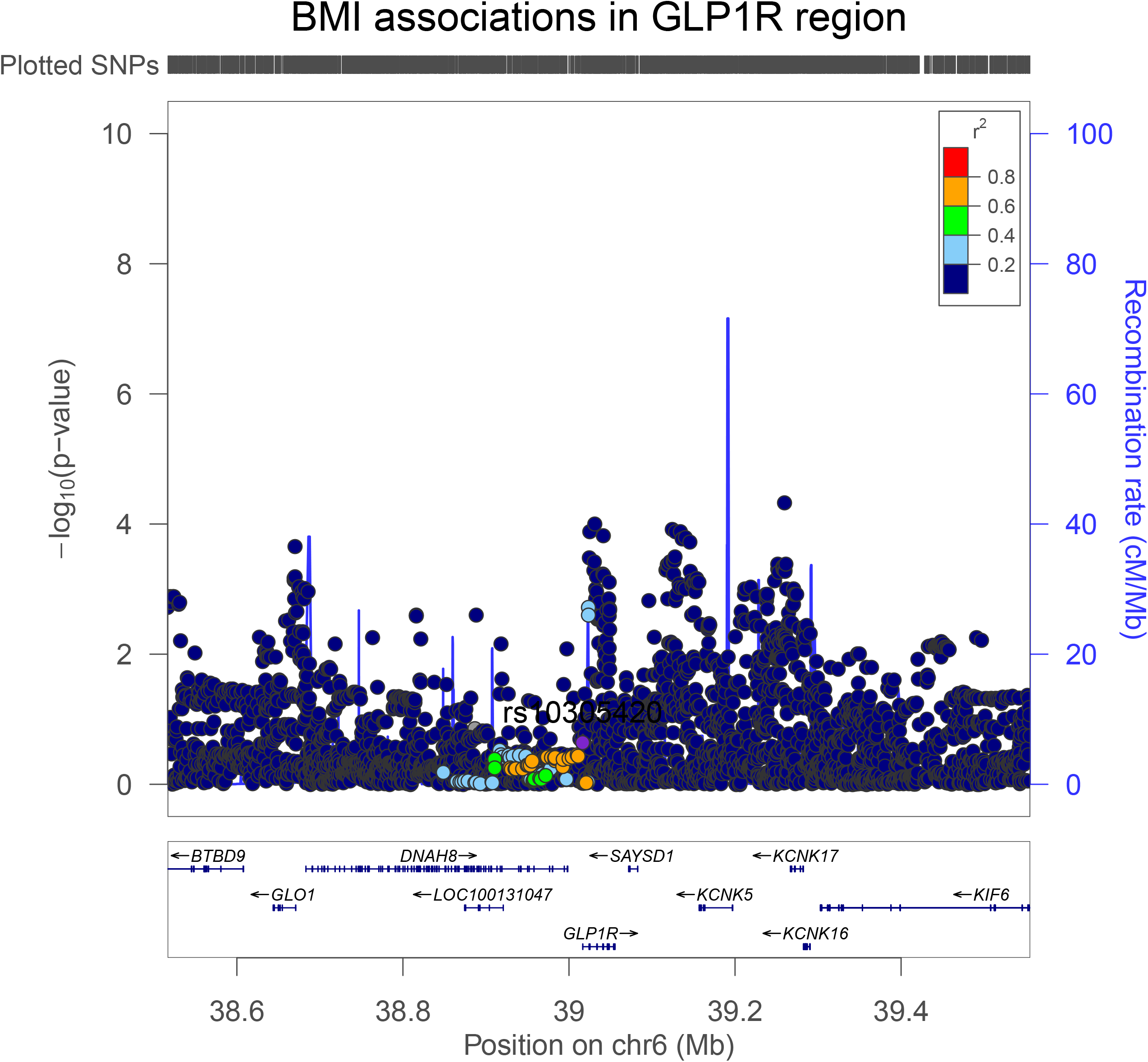

### Genetically-proxied PPARG perturbation and cancer risk

There was weak evidence for an association of genetically-proxied PPARG perturbation with an elevated risk of prostate cancer (OR 1.75, 95% CI 1.07-2.85; *P*=0.02), but little evidence of association with other cancer endpoints (**Table 2**). Findings for prostate cancer risk were consistent in iterative leave-one-out analysis (**Supplementary Table 7**). Colocalisation using marginal and conditional associations for T2D and prostate cancer in the *PPARG* locus suggested that T2D was unlikely to share a causal variant with this cancer in this region (posterior probability of a shared causal variant across models: ≤0.09%, posterior probability of distinct causal variants: ≤25%) (**Supplementary Table 8, Figure 8**).

**Table 2.** Mendelian randomization estimates examining the association of genetically-proxied perturbation of PPARG with site-specific and overall cancer risk

| **Outcome** | **N (cases, controls)** | **OR (95% CI)** | **P-value** |
| --- | --- | --- | --- |
| Breast cancer | 122,977; 105,974 | 0.67 (0.43-1.04) | 0.08 |
| ER+ Breast cancer | 69,501; 105,974 | 0.57 (0.38-0.85) | 6.45 x 10<sup>-3</sup> |
| ER- Breast cancer | 21,468; 105,974 | 1.14 (0.64-2.01) | 0.66 |
| Colorectal cancer | 58,221; 67,694 | 0.95 (0.51-1.75) | 0.86 |
| Colon cancer | 32,002; 64,159 | 1.22 (0.72-2.08) | 0.46 |
| Rectal cancer | 16,212; 64,159 | 0.82 (0.25-2.71) | 0.75 |
| Prostate cancer | 79,148; 61,106 | 1.75 (1.07-2.85) | 0.02 |
| Advanced prostate cancer | 15,167; 58,308 | 1.64 (0.62-4.33) | 0.32 |
| Overall cancer risk | 27,483; 372,016 | 0.72 (0.44-1.19) | 0.20 |

PPARG = peroxisome proliferator activated receptor gamma, ER+ = Oestrogen receptor positive, ER- = Oestrogen receptor negative. OR (95% CI) are scaled to represent the effect of genetically-proxied perturbation of PPARG equivalent to a 1 unit lowering of inverse rank-normal transformed HbA<sub>1c</sub> (mmol/mol). Advanced prostate cancer is defined as metastatic disease, Gleason score $\geq 8$ , prostate-specific antigen >100 or prostate cancer-related death.

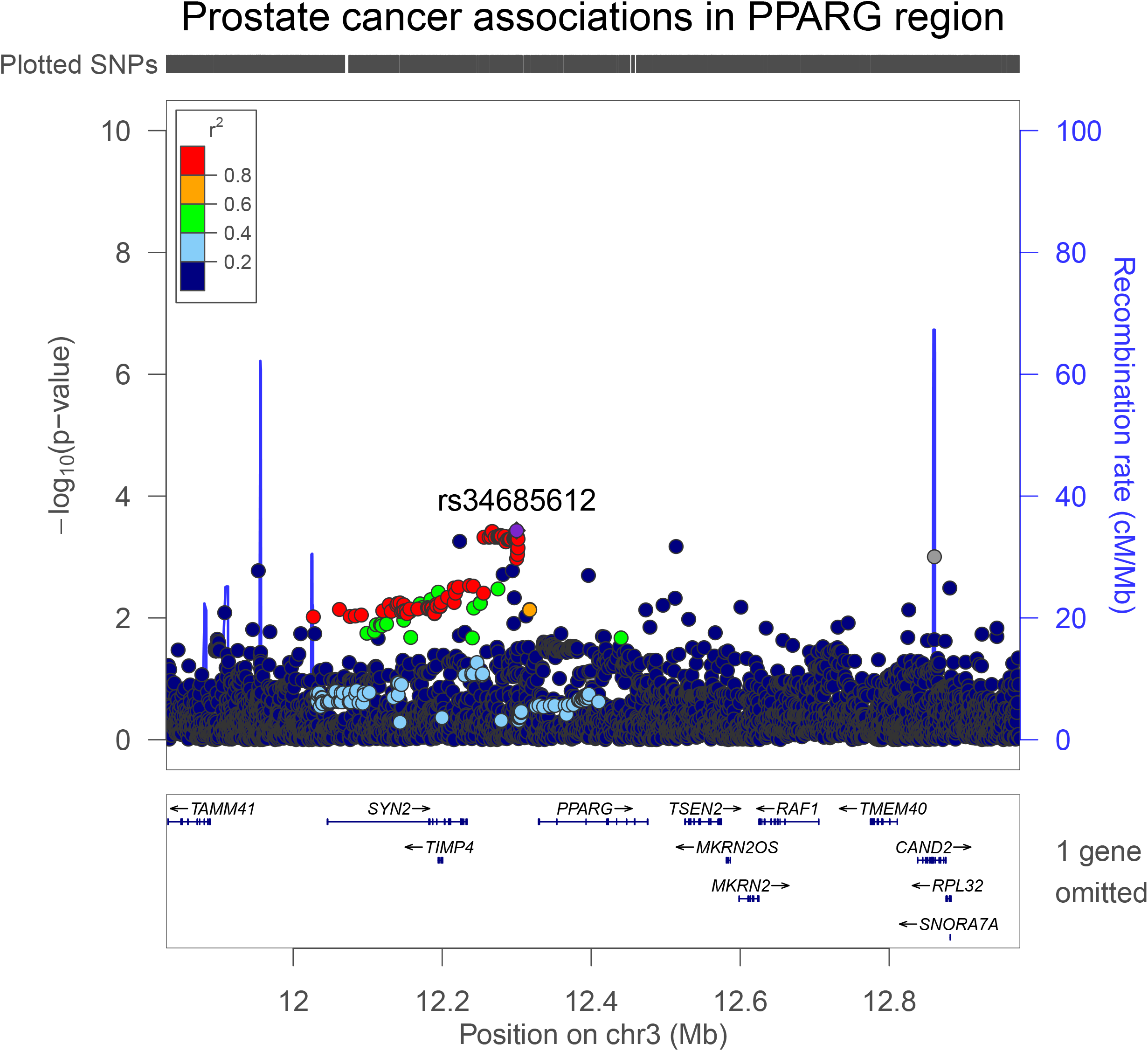

In subtype-stratified analyses, genetically-proxied PPARG perturbation was weakly associated with lower risk of ER+ breast cancer (OR 0.57, 95% CI 0.38-0.85; *P*=6.45 × 10^−3^). This finding was consistent in iterative leave-one-out analysis (**Supplementary Table 9**). Colocalisation using marginal and conditional associations for T2D and ER+ breast cancer in the *PPARG* locus reported a low posterior probability (H_4_<5%, posterior probability of distinct causal variants: ≤23%) of both traits sharing one or more causal variants within this region (**Supplementary Table 10, Figure 9**).

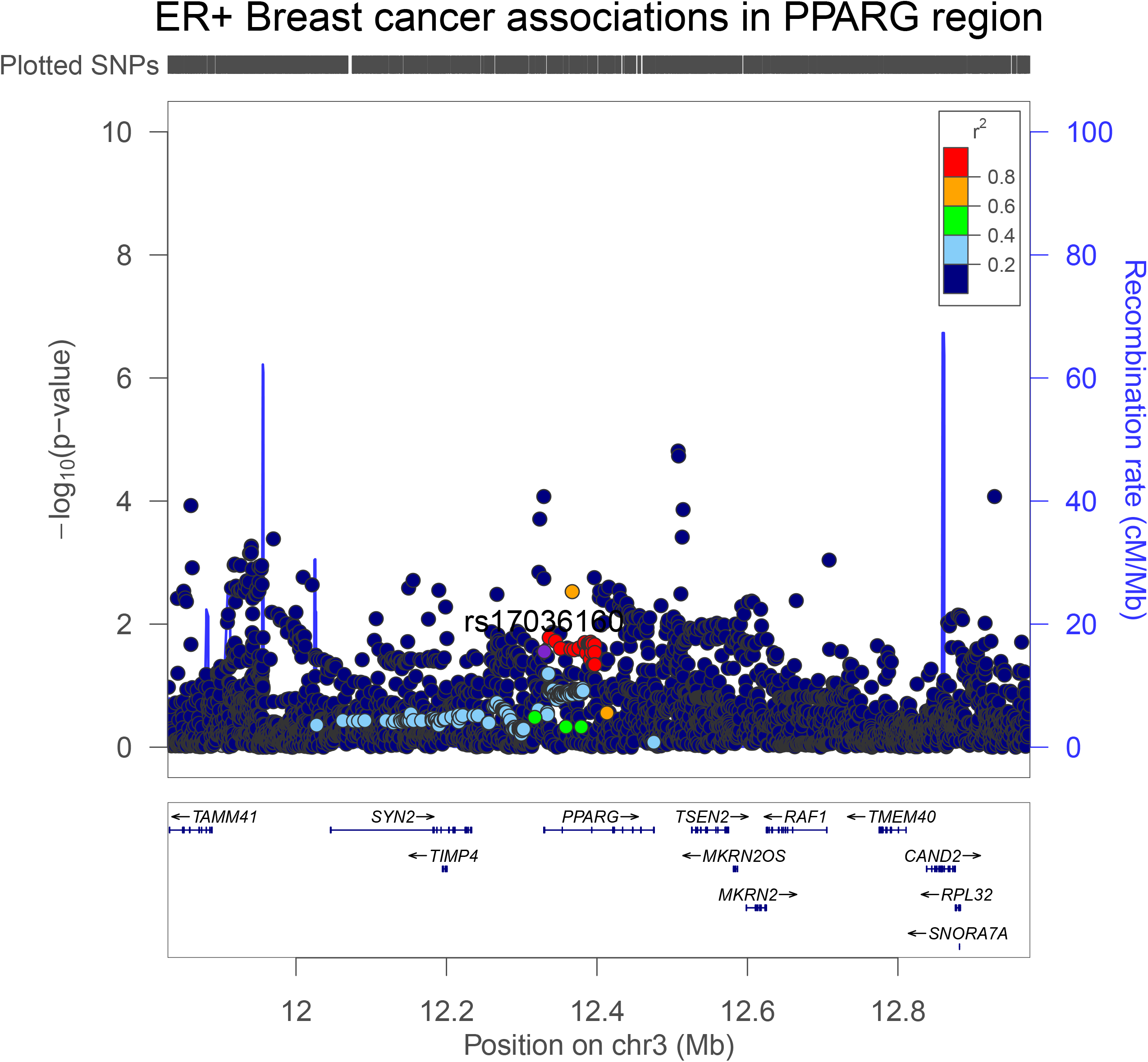

### Genetically-proxied ABCC8 and GLP1R perturbation and cancer risk

There was little Mendelian randomization evidence of association of genetically-proxied ABCC8 or GLP1R perturbation with site-specific or overall cancer risk (**Tables 3-4**).

**Table 3.** Mendelian randomization estimates examining the association of genetically-proxied perturbation of ABCC8 with site-specific and overall cancer risk

| **Outcome** | **N (cases, controls)** | **OR (95% CI)** | **P-value** |
| --- | --- | --- | --- |
| Breast cancer | 122,977; 105,974 | 2.09 (0.81-5.39) | 0.13 |
| ER+ Breast cancer | 69,501; 105,974 | 2.10 (0.66-6.74) | 0.21 |
| ER- Breast cancer | 21,468; 105,974 | 1.83 (0.48-6.97) | 0.38 |
| Colorectal cancer | 58,221; 67,694 | 0.61 (0.21-1.76) | 0.36 |
| Colon cancer | 32,002; 64,159 | 0.55 (0.15-1.93) | 0.35 |
| Rectal cancer | 16,212; 64,159 | 0.76 (0.15-3.87) | 0.74 |
| Prostate cancer | 79,148; 61,106 | 0.94 (0.37-2.43) | 0.91 |
| Advanced prostate cancer | 15,167; 58,308 | 1.67 (0.32-8.57) | 0.54 |
| Overall cancer risk | 27,483; 372,016 | 0.90 (0.31-2.59) | 0.85 |

ABCC8 = ATP Binding Cassette Subfamily C Member 8, ER+ = Oestrogen receptor positive, ER- = Oestrogen receptor negative. OR (95% CI) are scaled to represent the effect of genetically-proxied perturbation of ABCC8 equivalent to a 1 unit lowering of inverse rank-normal transformed HbA<sub>1c</sub> (mmol/mol). Advanced prostate cancer is defined as metastatic disease, Gleason score $\geq 8$ , prostate-specific antigen >100 or prostate cancer-related death.

**Table 4.** Mendelian randomization estimates examining the association of genetically-proxied perturbation of GLP1R with site-specific and overall cancer risk

| **Outcome** | **N (cases, controls)** | **OR (95% CI)** | **P-value** |
| --- | --- | --- | --- |
| Breast cancer | 122,977; 105,974 | 0.72 (0.33-1.58) | 0.42 |
| ER+ Breast cancer | 69,501; 105,974 | 0.81 (0.33-2.01) | 0.65 |
| ER- Breast cancer | 21,468; 105,974 | 0.48 (0.13-1.71) | 0.26 |
| Colorectal cancer | 58,221; 67,694 | 1.36 (0.50-3.68) | 0.55 |
| Colon cancer | 32,002; 64,159 | 1.94 (0.59-6.33) | 0.27 |
| Rectal cancer | 16,212; 64,159 | 1.13 (0.25-5.23) | 0.87 |
| Prostate cancer | 79,148; 61,106 | 0.87 (0.35-2.14) | 0.76 |
| Advanced prostate cancer | 15,167; 58,308 | 0.99 (0.10-9.51) | 0.99 |
| Overall cancer risk | 27,483; 372,016 | 1.21 (0.45-3.26) | 0.70 |

GLP1R = glucagon-like peptide 1 receptor, ER+ = Oestrogen receptor positive, ER- = Oestrogen receptor negative. OR (95% CI) are scaled to represent the effect of genetically-proxied perturbation of GLP1R equivalent to a 1 unit lowering of inverse rank-normal transformed HbA<sub>1c</sub> (mmol/mol). Advanced prostate cancer is defined as metastatic disease, Gleason score $\geq 8$ , prostate-specific antigen >100 or prostate cancer-related death.

## Discussion

In this Mendelian randomization analysis of up to 287,829 cases and 606,790 controls, we found weak evidence for an association of genetically-proxied PPARG perturbation with a higher risk of prostate cancer and lower risk of ER+ breast cancer. In colocalisation analysis, however, there was little evidence that genetic liability to type 2 diabetes and these cancer endpoints shared one or more causal variants within *PPARG*, though these analyses were likely underpowered given low posterior probabilities to support both H_3_ (i.e. distinct causal variants) and H_4_ (i.e. shared causal variants) across these analyses. We found little evidence of association of genetically-proxied GLP1R or ABCC8 perturbation with cancer risk.

Despite *in vivo* studies suggesting an important role of PPARG in prostate tumour growth and conventional epidemiological studies suggesting a link between pioglitazone use and elevated prostate cancer risk, our combined Mendelian randomization and colocalisation analyses did not find consistent evidence for an association of genetically-proxied PPARG perturbation with prostate cancer risk [6, 18]. Likewise, our findings are not consistent with some previous epidemiological studies which have reported links between rosiglitazone use and lower breast cancer risk and thiazolidinedione use and lower colorectal cancer risk [5, 19]. Though our analyses were powered to detect effect sizes comparable to those reported in some previous studies (e.g. a ∼60% increased prostate cancer risk among pioglitazone users and a ∼60% lower risk of colorectal cancer among thiazolidinedione users), they were likely less powered to detect other more modest effect sizes reported in the literature (e.g. ∼10% lower risk of breast cancer in rosiglitazone users)[19, 31, 56]. Interpretation of the pharmaco-epidemiological literature linking anti-diabetic medication use with cancer risk is challenging because of the likely susceptibility of many previous studies to residual confounding (e.g. by indication) due to the use of inappropriate comparator groups (i.e. non-medication users), the inclusion of “prevalent users” of medications in analyses, and the possibility of “immortal time” bias arising due to misalignment of the start of follow-up, eligibility, and treatment assignment of participants [21, 23].

Among the strengths of our analysis is the strict instrument selection and validation process employed. By using *cis*-acting variants, in close proximity to the genes that code for the drug targets of interest, horizontal pleiotropy should be minimised. In addition, we used strict positive control analysis (i.e. testing drug targets against established secondary effects of medications) and colocalisation analyses (including colocalisation analyses permitting multiple causal variants) to validate the selected instruments. Our use of a summary-data Mendelian randomization approach permitted us to leverage large-scale genetic data from several GWAS consortia, enhancing statistical power and precision of causal estimates.

There were several limitations to this analysis. First, we had sufficient statistical power to detect large effect sizes only and therefore cannot rule out more modest effects of the drug targets examined on cancer risk. Second, although colocalisation analyses of PPARG and cancer endpoints provided low posterior probabilities for shared causal variants, it should be noted that this may also reflect limited power. The low posterior probabilities supporting either shared or distinct causal variants across several colocalisation analyses suggests that many of these analyses may have been too underpowered to support either of these configurations evaluated. Third, the low posterior probability of shared causal variants in “positive control” colocalisation analyses for GLP1R and body mass index could reflect distinct signalling mechanisms influencing type 2 diabetes and body mass index in *GLP1R*, the presence of which would not necessarily influence the validity of this as an instrument for GLP1R signalling perturbation to affect glycaemic control [57]. Fourth, we were unable to evaluate the role of several anti-diabetic drug targets (i.e. DPP-4 and SGLT2) due to the absence of reliable genetic instruments for these targets. Fifth, our analyses were restricted to examining target-mediated (i.e. “on-target”) effects of anti-diabetic medications on cancer endpoints. Sixth, potential non-linear drug and/or time-dependent effects were not evaluated in the current analysis. Seventh, MR estimates reported represent long-term effects of target modulation in non-diabetic populations, whereas the clinical effects of these medications may be more pronounced among type 2 diabetes patients and could depend on length of medication use. Finally, samples were restricted to individuals of European ancestry and therefore the generalisability of these findings to non-European populations is unclear.

In conclusion, we developed novel instruments for PPARG, ABCC8, and GLP1R using strict validation protocols and evaluated the association of genetically-proxied perturbation of these targets with risk of cancer. In Mendelian randomization analysis we found weak evidence that genetically-proxied PPARG perturbation was associated with a higher risk of prostate cancer and a lower risk of ER+ breast cancer. There was little evidence of colocalisation for these findings, a necessary precondition to infer causality between PPARG perturbation and these cancer endpoints, which could reflect the absence of shared causal variants across type 2 diabetes liability and these cancer endpoints in *PPARG* or the low statistical power of these analyses. Further assessment of these drug targets using alternative molecular epidemiological approaches (e.g. using protein or expression quantitative trait loci or using direct circulating measures of these proteins) and/or studies using medical registry data (e.g. “target trial” analyses) may help to further corroborate findings presented in this analysis [58]. Finally, we found little evidence of an association of genetically-proxied ABCC8 and GLP1R perturbation with risk of breast, colorectal, prostate, or overall cancer risk.

## Data Availability

Summary genetic association data for select cancer endpoints were obtained from the public domain: breast cancer (https://bcac.ccge.medschl.cam.ac.uk/bcacdata/) and prostate cancer (http://practical.icr.ac.uk/). Summary genetic association data for colorectal cancer can be accessed by contacting GECCO (kafdem{at}fredhutch.org) and summary genetic association data for advanced prostate cancer can be accessed by contacting PRACTICAL (practical{at}icr.ac.uk). All other relevant data are within the manuscript and its Supporting Information files.

## Abbreviations

ABCC8: Sulfonylurea receptor 1
ALT: Alanine aminotransferase
AST: Aspartate aminotransferase
BCAC: Breast Cancer Association Consortium
BMI: Body mass index
CCFR: Colon Cancer Family Registry
CORECT: ColoRectal Transdisciplinary Study
CPRD: Clinical Practice Research Datalink
DPP-4: Dipeptidyl peptidase-4
GECCO: Genetics and Epidemiology of Colorectal Cancer Consortium
GLP1R: Glucacon-like peptide-1 receptor
GWAS: Genome-wide association study
HbA1c: Glycated haemoglobin
ICD: International Classification of Diseases
IVW: Inverse-variance weighted
MR: Mendelian randomization
PPARG: Peroxisome proliferator-activator nuclear receptor
PRACTICAL: Prostate Cancer Association Group to Investigate Cancer Associated Alterations in the Genome
SGLT2: Sodium-glucose cotransporter 2
SLC5A2: Sodium/glucose cotransporter 2 T2D Type 2 diabetes

## Funding

JY is supported by a Cancer Research UK Population Research Postdoctoral Fellowship (C68933/A28534). JY, EEV, SJL, RMM and KKT are supported by Cancer Research UK (C18281/A29019) programme grant (the Integrative Cancer Epidemiology Programme) (https://www.cancerresearchuk.org/). AC acknowledges funding from a Medical Research Council PhD studentship (MR/N013794/1). EEV is supported by a Diabetes UK RD Lawrence Fellowship (17/0005587). EEV and CJB are supported by Diabetes UK (17/0005587) and the World Cancer Research Fund (WCRF UK), as part of the World Cancer Research Fund International grant programme (IIG_2019_2009). MV is supported by I01-BX003362 from VA Office of R&D. RMM is a National Institute for Health Research Senior Investigator (NIHR202411). RMM is also supported by the NIHR Bristol Biomedical Research Centre which is funded by the NIHR (BRC-1215-20011) and is a partnership between University Hospitals Bristol and Weston NHS Foundation Trust and the University of Bristol. Department of Health and Social Care disclaimer: The views expressed are those of the author(s) and not necessarily those of the NHS, the NIHR or the Department of Health and Social Care. Disclaimer: Where authors are identified as personnel of the International Agency for Research on Cancer/World Health Organization, the authors alone are responsible for the views expressed in this article and they do not necessarily represent the decisions, policy or views of the International Agency for Research on Cancer/World Health Organization. This research is based on data from the Million Veteran Program, Office of Research and Development, Veterans Health Administration and was supported by award no. MVP000. This publication does not represent the views of the Department of Veterans Affairs, the US Food and Drug Administration, or the US Government. This research was also supported by funding from: the Department of Veterans Affairs awards I01-BX003362 (K.M.C). This publication does not represent the views of the Department of Veteran Affairs or the United States Government.

Genetics and Epidemiology of Colorectal Cancer Consortium (GECCO)-specific funding:

Genetics and Epidemiology of Colorectal Cancer Consortium (GECCO): National Cancer Institute, National Institutes of Health, U.S. Department of Health and Human Services (U01 CA137088, R01 CA059045, U01 CA164930, R21 CA191312, R01201407). Genotyping/Sequencing services were provided by the Center for Inherited Disease Research (CIDR) contract number HHSN268201700006I and HHSN268201200008I. This research was funded in part through the NIH/NCI Cancer Center Support Grant P30 CA015704. Scientific Computing Infrastructure at Fred Hutch funded by ORIP grant S10OD028685.

ASTERISK: a Hospital Clinical Research Program (PHRC-BRD09/C) from the University Hospital Center of Nantes (CHU de Nantes) and supported by the Regional Council of Pays de la Loire, the Groupement des Entreprises Françaises dans la Lutte contre le Cancer (GEFLUC), the Association Anne de Bretagne Génétique and the Ligue Régionale Contre le Cancer (LRCC).

The ATBC Study is supported by the Intramural Research Program of the U.S. National Cancer Institute, National Institutes of Health, Department of Health and Human Services.

CLUE II funding was from the National Cancer Institute (U01 CA86308, Early Detection Research Network; P30 CA006973), National Institute on Aging (U01 AG18033), and the American Institute for Cancer Research. The content of this publication does not necessarily reflect the views or policies of the Department of Health and Human Services, nor does mention of trade names, commercial products, or organizations imply endorsement by the US government.

Maryland Cancer Registry (MCR)

Cancer data was provided by the Maryland Cancer Registry, Center for Cancer Prevention and Control, Maryland Department of Health, with funding from the State of Maryland and the Maryland Cigarette Restitution Fund. The collection and availability of cancer registry data is also supported by the Cooperative Agreement NU58DP006333, funded by the Centers for Disease Control and Prevention. Its contents are solely the responsibility of the authors and do not necessarily represent the official views of the Centers for Disease Control and Prevention or the Department of Health and Human Services.

ColoCare: This work was supported by the National Institutes of Health (grant numbers R01 CA189184 (Li/Ulrich), U01 CA206110 (Ulrich/Li/Siegel/Figueiredo/Colditz, 2P30CA015704-40 (Gilliland), R01 CA207371 (Ulrich/Li)), the Matthias Lackas-Foundation, the German Consortium for Translational Cancer Research, and the EU TRANSCAN initiative.

The Colon Cancer Family Registry (CCFR, www.coloncfr.org) is supported in part by funding from the National Cancer Institute (NCI), National Institutes of Health (NIH) (award U01 CA167551). Support for case ascertainment was provided in part from the Surveillance, Epidemiology, and End Results (SEER) Program and the following U.S. state cancer registries: AZ, CO, MN, NC, NH; and by the Victoria Cancer Registry (Australia) and Ontario Cancer Registry (Canada). The CCFR Set-1 (Illumina 1M/1M-Duo) and Set-2 (Illumina Omni1-Quad) scans were supported by NIH awards U01 CA122839 and R01 CA143247 (to GC). The CCFR Set-3 (Affymetrix Axiom CORECT Set array) was supported by NIH award U19 CA148107 and R01 CA81488 (to SBG). The CCFR Set-4 (Illumina OncoArray 600K SNP array) was supported by NIH award U19 CA148107 (to SBG) and by the Center for Inherited Disease Research (CIDR), which is funded by the NIH to the Johns Hopkins University, contract number HHSN268201200008I. Additional funding for the OFCCR/ARCTIC was through award GL201-043 from the Ontario Research Fund (to BWZ), award 112746 from the Canadian Institutes of Health Research (to TJH), through a Cancer Risk Evaluation (CaRE) Program grant from the Canadian Cancer Society (to SG), and through generous support from the Ontario Ministry of Research and Innovation. The SFCCR Illumina HumanCytoSNP array was supported in part through NCI/NIH awards U01/U24 CA074794 and R01 CA076366 (to PAN). The content of this manuscript does not necessarily reflect the views or policies of the NCI, NIH or any of the collaborating centers in the Colon Cancer Family Registry (CCFR), nor does mention of trade names, commercial products, or organizations imply endorsement by the US Government, any cancer registry, or the CCFR.

COLON: The COLON study is sponsored by Wereld Kanker Onderzoek Fonds, including funds from grant 2014/1179 as part of the World Cancer Research Fund International Regular Grant Programme, by Alpe d’Huzes and the Dutch Cancer Society (UM 2012–5653, UW 2013-5927, UW2015-7946), and by TRANSCAN (JTC2012-MetaboCCC, JTC2013-FOCUS). The Nqplus study is sponsored by a ZonMW investment grant (98-10030); by PREVIEW, the project PREVention of diabetes through lifestyle intervention and population studies in Europe and around the World (PREVIEW) project which received funding from the European Union Seventh Framework Programme (FP7/2007–2013) under grant no. 312057; by funds from TI Food and Nutrition (cardiovascular health theme), a public– private partnership on precompetitive research in food and nutrition; and by FOODBALL, the Food Biomarker Alliance, a project from JPI Healthy Diet for a Healthy Life.

Colorectal Cancer Transdisciplinary (CORECT) Study: The CORECT Study was supported by the National Cancer Institute, National Institutes of Health (NCI/NIH), U.S. Department of Health and Human Services (grant numbers U19 CA148107, R01 CA081488, P30 CA014089, R01 CA197350; P01 CA196569; R01 CA201407; R01 CA242218),National Institutes of Environmental Health Sciences, National Institutes of Health (grant number T32 ES013678) and a generous gift from Daniel and Maryann Fong.

CORSA: The CORSA study was funded by Austrian Research Funding Agency (FFG) BRIDGE (grant 829675, to Andrea Gsur), the “Herzfelder’sche Familienstiftung” (grant to Andrea Gsur) and was supported by COST Action BM1206.

CPS-II: The American Cancer Society funds the creation, maintenance, and updating of the Cancer Prevention Study-II (CPS-II) cohort. This study was conducted with Institutional Review Board approval.

CRCGEN: Colorectal Cancer Genetics & Genomics, Spanish study was supported by Instituto de Salud Carlos III, co-funded by FEDER funds –a way to build Europe– (grants PI14-613 and PI09-1286), Agency for Management of University and Research Grants (AGAUR) of the Catalan Government (grant 2017SGR723), Junta de Castilla y León (grant LE22A10-2), the Spanish Association Against Cancer (AECC) Scientific Foundation grant GCTRA18022MORE and the Consortium for Biomedical Research in Epidemiology and Public Health (CIBERESP), action Genrisk. Sample collection of this work was supported by the Xarxa de Bancs de Tumors de Catalunya sponsored by Pla Director d’Oncología de Catalunya (XBTC), Plataforma Biobancos PT13/0010/0013 and ICOBIOBANC, sponsored by the Catalan Institute of Oncology. We thank CERCA Programme, Generalitat de Catalunya for institutional support.

Czech Republic CCS: This work was supported by the Grant Agency of the Czech Republic (18-09709S, 20-03997S), by the Grant Agency of the Ministry of Health of the Czech Republic (grants AZV NV18/03/00199 and AZV NV19-09-00237), and Charles University grants Unce/Med/006 and Progress Q28/LF1.

DACHS: This work was supported by the German Research Council (BR 1704/6-1, BR 1704/6-3, BR 1704/6-4, CH 117/1-1, HO 5117/2-1, HE 5998/2-1, KL 2354/3-1, RO 2270/8-1 and BR 1704/17-1), the Interdisciplinary Research Program of the National Center for Tumor Diseases (NCT), Germany, and the German Federal Ministry of Education and Research (01KH0404, 01ER0814, 01ER0815, 01ER1505A and 01ER1505B).

DALS: National Institutes of Health (R01 CA48998 to M. L. Slattery).

EDRN: This work is funded and supported by the NCI, EDRN Grant (U01 CA 84968-06).

EPIC: The coordination of EPIC is financially supported by International Agency for Research on Cancer (IARC) and also by the Department of Epidemiology and Biostatistics, School of Public Health, Imperial College London which has additional infrastructure support provided by the NIHR Imperial Biomedical Research Centre (BRC). The national cohorts are supported by: Danish Cancer Society (Denmark); Ligue Contre le Cancer, Institut Gustave Roussy, Mutuelle Générale de l’Education Nationale, Institut National de la Santé et de la Recherche Médicale (INSERM) (France); German Cancer Aid, German Cancer Research Center (DKFZ), German Institute of Human Nutrition Potsdam-Rehbruecke (DIfE), Federal Ministry of Education and Research (BMBF) (Germany); Associazione Italiana per la Ricerca sul Cancro-AIRC-Italy, Compagnia di SanPaolo and National Research Council (Italy); Dutch Ministry of Public Health, Welfare and Sports (VWS), Netherlands Cancer Registry (NKR), LK Research Funds, Dutch Prevention Funds, Dutch ZON (Zorg Onderzoek Nederland), World Cancer Research Fund (WCRF), Statistics Netherlands (The Netherlands); Health Research Fund (FIS) -Instituto de Salud Carlos III (ISCIII), Regional Governments of Andalucía, Asturias, Basque Country, Murcia and Navarra, and the Catalan Institute of Oncology -ICO (Spain); Swedish Cancer Society, Swedish Research Council and County Councils of Skåne and Västerbotten (Sweden); Cancer Research UK (14136 to EPIC-Norfolk; C8221/A29017 to EPIC-Oxford), Medical Research Council (1000143 to EPIC-Norfolk; MR/M012190/1 to EPIC-Oxford). (United Kingdom).

EPICOLON: This work was supported by grants from Fondo de Investigación Sanitaria/FEDER (PI08/0024, PI08/1276, PS09/02368, P111/00219, PI11/00681, PI14/00173, PI14/00230, PI17/00509, 17/00878, PI20/00113, PI20/00226, Acción Transversal de Cáncer), Xunta de Galicia (PGIDIT07PXIB9101209PR), Ministerio de Economia y Competitividad (SAF07-64873, SAF 2010-19273, SAF2014-54453R), Fundación Científica de la Asociación Española contra el Cáncer (GCB13131592CAST), Beca Grupo de Trabajo “Oncología” AEG (Asociación Española de Gastroenterología), Fundación Privada Olga Torres, FP7 CHIBCHA Consortium, Agència de Gestió d’Ajuts Universitaris i de Recerca (AGAUR, Generalitat de Catalunya, 2014SGR135, 2014SGR255, 2017SGR21, 2017SGR653), Catalan Tumour Bank Network (Pla Director d’Oncologia, Generalitat de Catalunya), PERIS (SLT002/16/00398, Generalitat de Catalunya), CERCA Programme (Generalitat de Catalunya) and COST Action BM1206 and CA17118. CIBERehd is funded by the Instituto de Salud Carlos III.

ESTHER/VERDI. This work was supported by grants from the Baden-Württemberg Ministry of Science, Research and Arts and the German Cancer Aid.

Harvard cohorts: HPFS is supported by the National Institutes of Health (P01 CA055075, UM1 CA167552, U01 CA167552, R01 CA137178, R01 CA151993, and R35 CA197735), NHS by the National Institutes of Health (P01 CA087969, UM1 CA186107, R01 CA137178, R01 CA151993, and R35 CA197735), and PHS by the National Institutes of Health (R01 CA042182).

Hawaii Adenoma Study: NCI grants R01 CA72520.

HCES-CRC: the Hwasun Cancer Epidemiology Study–Colon and Rectum Cancer (HCES-CRC; grants from Chonnam National University Hwasun Hospital, HCRI15011-1).

Kentucky: This work was supported by the following grant support: Clinical Investigator Award from Damon Runyon Cancer Research Foundation (CI-8); NCI R01CA136726.

LCCS: The Leeds Colorectal Cancer Study was funded by the Food Standards Agency and Cancer Research UK Programme Award (C588/A19167).

MCCS cohort recruitment was funded by VicHealth and Cancer Council Victoria. The MCCS was further supported by Australian NHMRC grants 509348, 209057, 251553 and 504711 and by infrastructure provided by Cancer Council Victoria. Cases and their vital status were ascertained through the Victorian Cancer Registry (VCR) and the Australian Institute of Health and Welfare (AIHW), including the National Death Index and the Australian Cancer Database.

MEC: National Institutes of Health (R37 CA54281, P01 CA033619, and R01 CA063464).

MECC: This work was supported by the National Institutes of Health, U.S. Department of Health and Human Services (R01 CA081488, R01 CA197350, U19 CA148107, R01 CA242218, and a generous gift from Daniel and Maryann Fong.

MSKCC: The work at Sloan Kettering in New York was supported by the Robert and Kate Niehaus Center for Inherited Cancer Genomics and the Romeo Milio Foundation. Moffitt: This work was supported by funding from the National Institutes of Health (grant numbers R01 CA189184, P30 CA076292), Florida Department of Health Bankhead-Coley Grant 09BN-13, and the University of South Florida Oehler Foundation. Moffitt contributions were supported in part by the Total Cancer Care Initiative, Collaborative Data Services Core, and Tissue Core at the H. Lee Moffitt Cancer Center & Research Institute, a National Cancer Institute-designated Comprehensive Cancer Center (grant number P30 CA076292).

NCCCS I & II: We acknowledge funding support for this project from the National Institutes of Health, R01 CA66635 and P30 DK034987.

NFCCR: This work was supported by an Interdisciplinary Health Research Team award from the Canadian Institutes of Health Research (CRT 43821); the National Institutes of Health, U.S. Department of Health and Human Serivces (U01 CA74783); and National Cancer Institute of Canada grants (18223 and 18226). The authors wish to acknowledge the contribution of Alexandre Belisle and the genotyping team of the McGill University and Génome Québec Innovation Centre, Montréal, Canada, for genotyping the Sequenom panel in the NFCCR samples. Funding was provided to Michael O. Woods by the Canadian Cancer Society Research Institute.

NSHDS: The research was supported by Biobank Sweden through funding from the Swedish Research Council (VR 2017-00650, VR 2017-01737), the Swedish Cancer Society (CAN 2017/581), Region Västerbotten (VLL-841671, VLL-833291), Knut and Alice Wallenberg Foundation (VLL-765961), and the Lion’s Cancer Research Foundation (several grants) and Insamlingsstiftelsen, both at Umeå University.

OSUMC: OCCPI funding was provided by Pelotonia and HNPCC funding was provided by the NCI (CA16058 and CA67941).

PLCO: Intramural Research Program of the Division of Cancer Epidemiology and Genetics and supported by contracts from the Division of Cancer Prevention, National Cancer Institute, NIH, DHHS. Funding was provided by National Institutes of Health (NIH), Genes, Environment and Health Initiative (GEI) Z01 CP 010200, NIH U01 HG004446, and NIH GEI U01 HG 004438.

SEARCH: The University of Cambridge has received salary support in respect of PDPP from the NHS in the East of England through the Clinical Academic Reserve. Cancer Research UK (C490/A16561); the UK National Institute for Health Research Biomedical Research Centres at the University of Cambridge.

SELECT: Research reported in this publication was supported in part by the National Cancer Institute of the National Institutes of Health under Award Numbers U10 CA37429 (CD Blanke), and UM1 CA182883 (CM Tangen/IM Thompson). The content is solely the responsibility of the authors and does not necessarily represent the official views of the National Institutes of Health.

SMS and REACH: This work was supported by the National Cancer Institute (grant P01 CA074184 to J.D.P. and P.A.N., grants R01 CA097325, R03 CA153323, and K05 CA152715 to P.A.N., and the National Center for Advancing Translational Sciences at the National Institutes of Health (grant KL2 TR000421 to A.N.B.-H.)

The Swedish Low-risk Colorectal Cancer Study: The study was supported by grants from the Swedish research council; K2015-55X-22674-01-4, K2008-55X-20157-03-3, K2006-72X-20157-01-2 and the Stockholm County Council (ALF project).

Swedish Mammography Cohort and Cohort of Swedish Men: This work is supported by the Swedish Research Council /Infrastructure grant, the Swedish Cancer Foundation, and the Karolinska Institute&s Distinguished Professor Award to Alicja Wolk.

UK Biobank: This research has been conducted using the UK Biobank Resource under Application Number 8614

VITAL: National Institutes of Health (K05 CA154337).

WHI: The WHI program is funded by the National Heart, Lung, and Blood Institute, National Institutes of Health, U.S. Department of Health and Human Services through contracts HHSN268201100046C, HHSN268201100001C, HHSN268201100002C, HHSN268201100003C, HHSN268201100004C, and HHSN271201100004C.

## Acknowledgments

The authors would like to thank the participants of the individual studies contributing to the Breast Cancer Association Consortium (BCAC) and the MAGIC (the Meta-Analyses of Glucose and Insulin-related traits Consortium) for their participation in these studies along with the principal investigators of these consortia for generating the data used for this analysis and for making these data available in the public domain.

Genetics and Epidemiology of Colorectal Cancer Consortium (GECCO)-specific acknowledgments: ASTERISK: We are very grateful to Dr. Bruno Buecher without whom this project would not have existed. We also thank all those who agreed to participate in this study, including the patients and the healthy control persons, as well as all the physicians, technicians and students.

CCFR: The Colon CFR graciously thanks the generous contributions of their study participants, dedication of study staff, and the financial support from the U.S. National Cancer Institute, without which this important registry would not exist. The authors would like to thank the study participants and staff of the Seattle Colon Cancer Family Registry and the Hormones and Colon Cancer study (CORE Studies).

CLUE II: We thank the participants of Clue II and appreciate the continued efforts of the staff at the Johns Hopkins George W. Comstock Center for Public Health Research and Prevention in the conduct of the Clue II Cohort Study.

COLON and NQplus: the authors would like to thank the COLON and NQplus investigators at Wageningen University & Research and the involved clinicians in the participating hospitals.

CORSA: We kindly thank all individuals who agreed to participate in the CORSA study. Furthermore, we thank all cooperating physicians and students and the Biobank Graz of the Medical University of Graz.

CPS-II: The authors thank the CPS-II participants and Study Management Group for their invaluable contributions to this research. The authors would also like to acknowledge the contribution to this study from central cancer registries supported through the Centers for Disease Control and Prevention National Program of Cancer Registries, and cancer registries supported by the National Cancer Institute Surveillance Epidemiology and End Results program.

Czech Republic CCS: We are thankful to all clinicians in major hospitals in the Czech Republic, without whom the study would not be practicable. We are also sincerely grateful to all patients participating in this study.

DACHS: We thank all participants and cooperating clinicians, and everyone who provided excellent technical assistance.

EDRN: We acknowledge all contributors to the development of the resource at University of Pittsburgh School of Medicine, Department of Gastroenterology, Department of Pathology, Hepatology and Nutrition and Biomedical Informatics.

EPIC: Where authors are identified as personnel of the International Agency for Research on Cancer/World Health Organization, the authors alone are responsible for the views expressed in this article and they do not necessarily represent the decisions, policy or views of the International Agency for Research on Cancer/World Health Organization.

EPICOLON: We are sincerely grateful to all patients participating in this study who were recruited as part of the EPICOLON project. We acknowledge the Spanish National DNA Bank, Biobank of Hospital Clínic–IDIBAPS and Biobanco Vasco for the availability of the samples. The work was carried out (in part) at the Esther Koplowitz Centre, Barcelona.

Harvard cohorts (HPFS, NHS, PHS): The study protocol was approved by the institutional review boards of the Brigham and Women’s Hospital and Harvard T.H. Chan School of Public Health, and those of participating registries as required. We acknowledge Channing Division of Network Medicine, Department of Medicine, Brigham and Women‘s Hospital as home of the NHS. The authors would like to acknowledge the contribution to this study from central cancer registries supported through the Centers for Disease Control and Prevention’s National Program of Cancer Registries (NPCR) and/or the National Cancer Institute’s Surveillance, Epidemiology, and End Results (SEER) Program. Central registries may also be supported by state agencies, universities, and cancer centers. Participating central cancer registries include the following: Alabama, Alaska, Arizona, Arkansas, California, Colorado, Connecticut, Delaware, Florida, Georgia, Hawaii, Idaho, Indiana, Iowa, Kentucky, Louisiana, Massachusetts, Maine, Maryland, Michigan, Mississippi, Montana, Nebraska, Nevada, New Hampshire, New Jersey, New Mexico, New York, North Carolina, North Dakota, Ohio, Oklahoma, Oregon, Pennsylvania, Puerto Rico, Rhode Island, Seattle SEER Registry, South Carolina, Tennessee, Texas, Utah, Virginia, West Virginia, Wyoming. The authors assume full responsibility for analyses and interpretation of these data.

Kentucky: We would like to acknowledge the staff at the Kentucky Cancer Registry.

LCCS: We acknowledge the contributions of Jennifer Barrett, Robin Waxman, Gillian Smith and Emma Northwood in conducting this study.

NCCCS I & II: We would like to thank the study participants, and the NC Colorectal Cancer Study staff.

NSHDS investigators thank the Västerbotten Intervention Programme, the Northern Sweden MONICA study, the Biobank Research Unit at Umeå University and Biobanken Norr at Region Västerbotten for providing data and samples and acknowledge the contribution from Biobank Sweden, supported by the Swedish Research Council.

PLCO: The authors thank the PLCO Cancer Screening Trial screening center investigators and the staff from Information Management Services Inc and Westat Inc. Most importantly, we thank the study participants for their contributions that made this study possible.

Cancer incidence data have been provided by the District of Columbia Cancer Registry, Georgia Cancer Registry, Hawaii Cancer Registry, Minnesota Cancer Surveillance System, Missouri Cancer Registry, Nevada Central Cancer Registry, Pennsylvania Cancer Registry, Texas Cancer Registry, Virginia Cancer Registry, and Wisconsin Cancer Reporting System. All are supported in part by funds from the Center for Disease Control and Prevention, National Program for Central Registries, local states or by the National Cancer Institute, Surveillance, Epidemiology, and End Results program. The results reported here and the conclusions derived are the sole responsibility of the authors.

SEARCH: We thank the SEARCH team

SELECT: We thank the research and clinical staff at the sites that participated on SELECT study, without whom the trial would not have been successful. We are also grateful to the 35,533 dedicated men who participated in SELECT.

WHI: The authors thank the WHI investigators and staff for their dedication, and the study participants for making the program possible. A full listing of WHI investigators can be found at: http://www.whi.org/researchers/Documents%20%20Write%20a%20Paper/WHI%20Investigator%20Short%20List.pdf

