## Supplementary Material for "Genetically-proxied anti-diabetic drug target perturbation and risk of cancer: a Mendelian randomization analysis"

Supplementary Table 1. R^2^ and F-statistics for instruments used to proxy PPARG, ABCC8, and GLP1R perturbation

| **Target** | **R^2^** | **F-statistic** |
| --- | --- | --- |
| PPARG | 0.00070 -0.00119 | 285.82-487.14 |
| ABCC8 | 0.00014-0.00027 | 59.11-111.98 |
| GLP1R | 0.00014-0.00022 | 56.32-88.82 |

Range of R^2^ and F-statistics correspond to estimates of these metrics across instruments constructed using independent (r^2^<0.001) and weakly correlated (r^2^<0.20) SNPs.

Supplementary Table 2. Estimates of the smallest detectable OR per unit change in drug target-mediated inverse rank-normal transformed HbA_1c_ reduction (mmol/mol) where there is 80% power to detect an effect (α=0.05)

| **Cancer endpoint** | **PPARG** | **ABCC8** | **GLP1R** |
| --- | --- | --- | --- |
| Breast cancer | 1.40-1.55 | 2.03-2.65 | 2.22-2.72 |
| ER+ Breast cancer | 1.49-1.68 | 2.28-3.11 | 2.53-3.20 |
| ER- Breast cancer | 1.83-2.21 | 3.55-5.71 | 4.14-5.96 |
| Colorectal cancer | 1.58-1.82 | 2.60-3.73 | 2.92-3.85 |
| Colon cancer | 1.74-2.06 | 3.18-4.92 | 3.67-5.11 |
| Rectal cancer | 2.00-2.54 | 4.42-7.73 | 5.31-8.13 |
| Prostate cancer | 1.55-1.77 | 2.49-3.50 | 2.78-3.61 |
| Advanced prostate cancer | 2.09-2.62 | 4.67-8.34 | 5.64-8.78 |
| Overall cancer risk | 1.66-1.94 | 2.88-4.28 | 3.28-4.44 |

Ranges correspond to differences in power estimates across instruments constructed using independent (r^2^<0.001) and weakly correlated (r^2^<0.20) SNPs.

Supplementary Table 3. Posterior probabilities under differing hypotheses relating the associations between T2D variants in or within proximity to the *PPARG* locus and ALT levels

| **Configuration** | **H_0_** | **H_1_** | **H_2_** | **H_3_** | **H_4_** |
| --- | --- | --- | --- | --- | --- |
|  | 8.33 x 10^-63^ | 9.24 x 10^-23^ | 7.43 x 10^-42^ | 8.15 x 10^-2^ | 0.92 |

H_0_ = neither T2D nor ALT has a genetic association in the region, H_1_ = only T2D has a genetic association in the region, H_2_ = only ALT has a genetic association in the region, H_3_ = both T2D and ALT are associated but have different causal variants, H_4_= both T2D and ALT are associated and share a single causal variant.

Supplementary Table 4. Posterior probabilities under differing hypotheses relating the associations between T2D variants in or within proximity to the *PPARG* locus and AST levels

| **Configuration** | **H_0_** | **H_1_** | **H_2_** | **H_3_** | **H_4_** |
| --- | --- | --- | --- | --- | --- |
|  | 1.58 x 10^-50^ | 1.76 x 10^-10^ | 1.46 x 10^-41^ | 0.16 | 0.84 |

H_0_ = neither T2D nor AST has a genetic association in the region, H_1_ = only T2D has a genetic association in the region, H_2_ = only AST has a genetic association in the region, H_3_ = both T2D and AST are associated but have different causal variants, H_4_= both T2D and AST are associated and share a single causal variant.

Supplementary Table 5. Posterior probabilities under differing hypotheses relating the associations between T2D variants in or within proximity to the *ABCC8* locus and BMI levels

| **Configuration** | **H_0_** | **H_1_** | **H_2_** | **H_3_** | **H_4_** |
| --- | --- | --- | --- | --- | --- |
|  | 8.07 x 10^-43^ | 2.70 x 10^-2^ | 1.09 x 10^-42^ | 3.56 x 10^-2^ | 0.94 |

H_0_ = neither T2D nor BMI has a genetic association in the region, H_1_ = only T2D has a genetic association in the region, H_2_ = only BMI has a genetic association in the region, H_3_ = both T2D and BMI are associated but have different causal variants, H_4_= both T2D and BMI are associated and share a single causal variant.

Supplementary Table 6. Posterior probabilities under differing hypotheses relating the associations between T2D in or within proximity to the *GLP1R* locus and BMI levels

| **T2D SNP** | **BMI SNP** | **H_0_** | **H_1_** | **H_2_** | **H_3_** | **H_4_** |
| --- | --- | --- | --- | --- | --- | --- |
| Marginal | Marginal | 2.21 x 10^-5^ | 0.88 | 2.91 x 10^-6^ | 0.12 | 2.27 x 10^-3^ |
| rs10305420* | Marginal | 2.40 x 10^-6^ | 0.89 | 2.91 x 10^-7^ | 0.11 | 2.16 x 10^-3^ |
| rs147718699^†^ | Marginal | 5.15 x 10^-5^ | 0.89 | 6.31 x 10^-6^ | 0.11 | 3.08 x10^-3^ |
| rs2115200^†^ | Marginal | 4.86 x 10^-5^ | 0.89 | 5.90 x 10^-6^ | 0.11 | 2.94 x 10^-3^ |
| rs34179517^†^ | Marginal | 1.66 x 10^-7^ | 0.89 | 2.01 x 10^-8^ | 0.11 | 4.93 x 10^-3^ |

Marginal = SNP associations that are unconditioned (on either the sentinel SNP or additional conditionally independent genome-wide significant SNP for each respective trait), * = Sentinel SNP, ^†^ = Conditionally independent and significant (P<5x10^-8^) SNP, H_0_ = neither T2D nor BMI has a genetic association in the region, H_1_ = only T2D has a genetic association in the region, H_2_ = only BMI has a genetic association in the region, H_3_ = both T2D and BMI are associated but have different causal variants, H_4_= both T2D and BMI are associated and share a single causal variant.

Supplementary Table 7. Iterative leave-one-out analysis for Mendelian randomization analysis of genetically-proxied PPARG perturbation and risk of prostate cancer

| **SNP removed** | **OR (95% CI)** |
| --- | --- |
| rs143888770 | 1.74 (1.02-2.97) |
| rs150535373 | 1.75 (1.03-2.99) |
| rs17819328 | 1.81 (1.02-3.24) |
| rs4135247 | 1.51 (0.70-3.29) |
| rs4135300 | 1.81 (1.08-3.04) |
| rs598747 | 1.87 (1.19-2.94) |
| rs7637403 | 1.77 (1.18-2.66) |

OR (95% CI) are scaled to represent the effect of genetically-proxied perturbation of PPARG equivalent to a 1 unit decrease in IRNT HbA_1c_.

Supplementary Table 8. Posterior probabilities under differing hypotheses relating the associations between T2D in or within proximity to the *PPARG* locus and prostate cancer risk

| **T2D SNP** | **PCa SNP** | **H_0_** | **H_1_** | **H_2_** | **H_3_** | **H_4_** |
| --- | --- | --- | --- | --- | --- | --- |
| Marginal | Marginal | 6.68 x 10^-41^ | 0.74 | 2.26 x 10^-41^ | 0.25 | 8.14 x 10^-3^ |
| rs17036160* | Marginal | 3.84 x 10^-51^ | 0.81 | 8.80 x 10^-52^ | 0.18 | 8.87 x 10^-3^ |
| rs7650482^†^ | Marginal | 7.79 x 10^-41^ | 0.81 | 1.78 x 10^-41^ | 0.18 | 8.89 x 10^-3^ |
| rs9872031^†^ | Marginal | 7.79 x 10^-41^ | 0.81 | 1.78 x 10^-41^ | 0.18 | 8.89 x 10^-3^ |

Marginal = SNP associations that are unconditioned (on either the sentinel SNP or additional conditionally independent genome-wide significant SNP for each respective trait), * = Sentinel SNP, ^†^ = Conditionally independent and significant (*P*<5x10^-8^) SNP, H_0_ = neither T2D nor prostate cancer risk has a genetic association in the region, H_1_ = only T2D has a genetic association in the region, H_2_ = only prostate cancer risk has a genetic association in the region, H_3_ = both T2D and prostate cancer risk are associated but have different causal variants, H_4_= both T2D and prostate cancer risk are associated and share a single causal variant.

Supplementary Table 9. Iterative leave-one-out analysis for Mendelian randomization analysis of genetically-proxied PPARG perturbation and risk of ER+ breast cancer

| **SNP removed** | **OR (95% CI)** |
| --- | --- |
| rs143888770 | 0.58 (0.38-0.87) |
| rs150535373 | 0.56 (0.38-0.82) |
| rs17819328 | 0.53 (0.34-0.83) |
| rs4135247 | 0.63 (0.33-1.19) |
| rs4135300 | 0.55 (0.37-0.83) |
| rs598747 | 0.57 (0.36-0.89) |
| rs7637403 | 0.57 (0.38-0.88) |

OR (95% CI) are scaled to represent the effect of genetically-proxied perturbation of PPARG equivalent to a unit decrease in IRNT HbA_1c_.

Supplementary Table 10. Posterior probabilities under differing hypotheses relating the associations between T2D in or within proximity to the *PPARG* locus and ER+ breast cancer risk

| **T2D SNP** | **BCa SNP** | **H_0_** | **H_1_** | **H_2_** | **H_3_** | **H_4_** |
| --- | --- | --- | --- | --- | --- | --- |
| Marginal | Marginal | 6.50 x 10^-41^ | 0.72 | 2.08 x 10^-41^ | 0.23 | 4.74 x 10^-2^ |
| rs17036160* | Marginal | 3.60 x 10^-51^ | 0.76 | 9.29 x 10^-52^ | 0.20 | 4.92 x 10^-2^ |
| rs7650482^†^ | Marginal | 7.32 x 10^-41^ | 0.76 | 1.88 x 10^-41^ | 0.19 | 4.81 x 10^-2^ |
| rs9872031^†^ | Marginal | 7.32 x 10^-41^ | 0.76 | 1.88 x 10^-41^ | 0.19 | 4.81 x 10^-2^ |

Marginal = SNP associations that are unconditioned (on either the sentinel SNP or additional conditionally independent genome-wide significant SNP for each respective trait), * = Sentinel SNP, ^†^ = Conditionally independent and significant (*P*<5x10^-8^) SNP, H_0_ = neither T2D nor ER+ breast cancer risk has a genetic association in the region, H_1_ = only T2D has a genetic association in the region, H_2_ = only ER+ breast cancer risk has a genetic association in the region, H_3_ = both T2D and ER+ breast cancer risk are associated but have different causal variants, H_4_= both T2D and ER+ breast cancer risk are associated and share a single causal variant.

**The PRACTICAL Consortium**

**<http://practical.icr.ac.uk/>**

Rosalind A. Eeles^1,2^, Christopher A. Haiman^3^, Zsofia Kote-Jarai^1^, Fredrick R. Schumacher^4,5^, Sara Benlloch^6,1^, Ali Amin Al Olama^6,7^, Kenneth Muir^8,9^, Sonja I. Berndt^10^, David V. Conti^3^, Fredrik Wiklund^11^, Stephen Chanock^10^, Ying Wang^12^, Victoria L. Stevens^12^, Catherine M. Tangen^13^, Jyotsna Batra^14,15^, Judith A. Clements^14,15^, APCB BioResource (Australian Prostate Cancer BioResource)^14,15^, Henrik Grönberg^11^, Nora Pashayan^16,17^, Johanna Schleutker^18,19^, Demetrius Albanes^10^, Stephanie Weinstein^10^, Alicja Wolk^20^, Catharine M. L. West^21^, Lorelei A. Mucci^22^, Géraldine Cancel-Tassin^23,24^, Stella Koutros^10^, Karina Dalsgaard Sørensen^25,26^, Eli Marie Grindedal^27^, David E. Neal^28,29,30^, Freddie C. Hamdy^31,32^, Jenny L. Donovan^33^, Ruth C. Travis^34^, Robert J. Hamilton^35,36^, Sue Ann Ingles^37^, Barry S. Rosenstein^38,39^, Yong-Jie Lu^40^, Graham G. Giles^41,42,43^, Adam S. Kibel^44^, Ana Vega^45,46,47^, Manolis Kogevinas^48,49,50,51^, Kathryn L. Penney^52^, Jong Y. Park^53^, Janet L. Stanford^54,55^, Cezary Cybulski^56^, Børge G. Nordestgaard^57,58^, Sune F. Nielsen^57,58^, Hermann Brenner^59,60,61^, Christiane Maier^62^, Jeri Kim^63^, Esther M. John^64^, Manuel R. Teixeira^65,66,67^, Susan L. Neuhausen^68^, Kim De Ruyck^69^, Azad Razack^70^, Lisa F. Newcomb^54,71^, Davor Lessel^72^, Radka Kaneva^73^, Nawaid Usmani^74,75^, Frank Claessens^76^, Paul A. Townsend^77,78^, Jose Esteban Castelao^79^, Monique J. Roobol^80^, Florence Menegaux^81^, Kay-Tee Khaw^82^, Lisa Cannon-Albright^83,84^, Hardev Pandha^78^, Stephen N. Thibodeau^85^, David J. Hunter^86^, Peter Kraft^87^, William J. Blot^88,89^, Elio Riboli^90^

1 The Institute of Cancer Research, London, SM2 5NG, UK

2 Royal Marsden NHS Foundation Trust, London, SW3 6JJ, UK

3 Center for Genetic Epidemiology, Department of Preventive Medicine, Keck School of Medicine, University of Southern California/Norris Comprehensive Cancer Center, Los Angeles, CA 90015, USA

4 Department of Population and Quantitative Health Sciences, Case Western Reserve University, Cleveland, OH 44106–7219, USA

5 Seidman Cancer Center, University Hospitals, Cleveland, OH 44106, USA

6 Centre for Cancer Genetic Epidemiology, Department of Public Health and Primary Care, University of Cambridge, Strangeways Research Laboratory, Cambridge CB1 8RN, UK

7 University of Cambridge, Department of Clinical Neurosciences, Stroke Research Group, R3, Box 83, Cambridge Biomedical Campus, Cambridge CB2 0QQ, UK

8 Division of Population Health, Health Services Research and Primary Care, University of Manchester, Oxford Road, Manchester, M13 9PL, UK

9 Warwick Medical School, University of Warwick, Coventry, CV4 7AL, UK

10 Division of Cancer Epidemiology and Genetics, National Cancer Institute, NIH, Bethesda, Maryland, 20892, USA

11 Department of Medical Epidemiology and Biostatistics, Karolinska Institute, SE-171 77 Stockholm, Sweden

12 Department of Population Science, American Cancer Society, 250 Williams Street, Atlanta, GA 30303, USA

13 SWOG Statistical Center, Fred Hutchinson Cancer Research Center, Seattle, WA 98109, USA

14 Australian Prostate Cancer Research Centre-Qld, Institute of Health and Biomedical Innovation and School of Biomedical Sciences, Queensland University of Technology, Brisbane QLD 4059, Australia

15 Translational Research Institute, Brisbane, Queensland 4102, Australia

16 Department of Applied Health Research, University College London, London, WC1E 7HB, UK

17 Centre for Cancer Genetic Epidemiology, Department of Oncology, University of Cambridge, Strangeways Laboratory, Worts Causeway, Cambridge, CB1 8RN, UK

18Institute of Biomedicine, University of Turku, Finland

19 Department of Medical Genetics, Genomics, Laboratory Division, Turku University Hospital, PO Box 52, 20521 Turku, Finland

20 Department of Surgical Sciences, Uppsala University, 75185 Uppsala, Sweden

21 Division of Cancer Sciences, University of Manchester, Manchester Academic Health Science Centre, Radiotherapy Related Research, The Christie Hospital NHS Foundation Trust, Manchester, M13 9PL UK

22 Department of Epidemiology, Harvard T. H. Chan School of Public Health, Boston, MA 02115, USA

23 CeRePP, Tenon Hospital, F-75020 Paris, France

24 Sorbonne Universite, GRC n°5, AP-HP, Tenon Hospital, 4 rue de la Chine, F-75020 Paris, France

25 Department of Molecular Medicine, Aarhus University Hospital, Palle Juul-Jensen Boulevard 99, 8200 Aarhus N, Denmark

26 Department of Clinical Medicine, Aarhus University, DK-8200 Aarhus N

27 Department of Medical Genetics, Oslo University Hospital, 0424 Oslo, Norway

28 Nuffield Department of Surgical Sciences, University of Oxford, Room 6603, Level 6, John Radcliffe Hospital, Headley Way, Headington, Oxford, OX3 9DU, UK

29 University of Cambridge, Department of Oncology, Box 279, Addenbrooke’s Hospital, Hills Road, Cambridge CB2 0QQ, UK

30 Cancer Research UK, Cambridge Research Institute, Li Ka Shing Centre, Cambridge, CB2 0RE, UK

31 Nuffield Department of Surgical Sciences, University of Oxford, Oxford, OX1 2JD, UK

32 Faculty of Medical Science, University of Oxford, John Radcliffe Hospital, Oxford, UK

33 Population Health Sciences, Bristol Medical School, University of Bristol, BS8 2PS, UK

34 Cancer Epidemiology Unit, Nuffield Department of Population Health, University of Oxford, Oxford, OX3 7LF, UK

35 Dept. of Surgical Oncology, Princess Margaret Cancer Centre, Toronto ON M5G 2M9, Canada

36 Dept. of Surgery (Urology), University of Toronto, Canada

37 Department of Preventive Medicine, Keck School of Medicine, University of Southern California/Norris Comprehensive Cancer Center, Los Angeles, CA 90015, USA

38 Department of Radiation Oncology and Department of Genetics and Genomic Sciences, Box 1236, Icahn School of Medicine at Mount Sinai, One Gustave L. Levy Place, New York, NY 10029, USA

39 Department of Genetics and Genomic Sciences, Icahn School of Medicine at Mount Sinai, New York, NY 10029–5674, USA

40 Centre for Cancer Biomarker and Biotherapeutics, Barts Cancer Institute, Queen Mary University of London, John Vane Science Centre, Charterhouse Square, London, EC1M 6BQ, UK

41 Cancer Epidemiology Division, Cancer Council Victoria, 615 St Kilda Road, Melbourne, VIC 3004, Australia

42 Centre for Epidemiology and Biostatistics, Melbourne School of Population and Global Health, The University of Melbourne, Grattan Street, Parkville, VIC 3010, Australia

43 Precision Medicine, School of Clinical Sciences at Monash Health, Monash University, Clayton, Victoria 3168, Australia

44 Division of Urologic Surgery, Brigham and Womens Hospital, 75 Francis Street, Boston, MA 02115, USA

45 Fundación Pública Galega Medicina Xenómica, Santiago de Compostela, 15706, Spain

46 Instituto de Investigación Sanitaria de Santiago de Compostela, Santiago De Compostela, 15706, Spain

47 Centro de Investigación en Red de Enfermedades Raras (CIBERER), Spain

48 ISGlobal, Barcelona, Spain

49 IMIM (Hospital del Mar Medical Research Institute), Barcelona, Spain

50 CIBER Epidemiología y Salud Pública (CIBERESP), 28029 Madrid, Spain

51 Universitat Pompeu Fabra (UPF), Barcelona, Spain

52 Channing Division of Network Medicine, Department of Medicine, Brigham and Women’s Hospital/Harvard Medical School, Boston, MA 02115, USA

53 Department of Cancer Epidemiology, Moffitt Cancer Center, 12902 Magnolia Drive, Tampa, FL 33612, USA

54 Division of Public Health Sciences, Fred Hutchinson Cancer Research Center, Seattle, Washington, 98109–1024, USA

55 Department of Epidemiology, School of Public Health, University of Washington, Seattle, Washington 98195, USA

56 International Hereditary Cancer Center, Department of Genetics and Pathology, Pomeranian Medical University, 70–115 Szczecin, Poland

57 Faculty of Health and Medical Sciences, University of Copenhagen, 2200 Copenhagen, Denmark

58 Department of Clinical Biochemistry, Herlev and Gentofte Hospital, Copenhagen University Hospital, Herlev, 2200 Copenhagen, Denmark

59 Division of Clinical Epidemiology and Aging Research, German Cancer Research Center (DKFZ), D-69120, Heidelberg, Germany

60 German Cancer Consortium (DKTK), German Cancer Research Center (DKFZ), D-69120 Heidelberg, Germany

61 Division of Preventive Oncology, German Cancer Research Center (DKFZ) and National Center for Tumor Diseases (NCT), Im Neuenheimer Feld 460, 69120 Heidelberg, Germany

62 Humangenetik Tuebingen, Paul-Ehrlich-Str 23, D-72076 Tuebingen, Germany

63 The University of Texas M. D. Anderson Cancer Center, Department of Genitourinary Medical Oncology, 1515 Holcombe Blvd., Houston, TX 77030, USA

64 Departments of Epidemiology & Population Health and of Medicine, Division of Oncology, Stanford Cancer Institute, Stanford University School of Medicine, Stanford, CA 94304 USA

65 Department of Genetics, Portuguese Oncology Institute of Porto (IPO-Porto), 4200–072 Porto, Portugal

66 Biomedical Sciences Institute (ICBAS), University of Porto, 4050–313 Porto, Portugal

67 Cancer Genetics Group, IPO-Porto Research Center (CI-IPOP), Portuguese Oncology Institute of Porto (IPO-Porto), 4200–072 Porto, Portugal

68 Department of Population Sciences, Beckman Research Institute of the City of Hope, 1500 East Duarte Road, Duarte, CA 91010, 626-256-HOPE (4673)

69 Ghent University, Faculty of Medicine and Health Sciences, Basic Medical Sciences, Proeftuinstraat 86, B-9000 Gent

70 Department of Surgery, Faculty of Medicine, University of Malaya, 50603 Kuala Lumpur, Malaysia

71 Department of Urology, University of Washington, 1959 NE Pacific Street, Box 356510, Seattle, WA 98195, USA

72 Institute of Human Genetics, University Medical Center Hamburg-Eppendorf, D-20246 Hamburg, Germany

73 Molecular Medicine Center, Department of Medical Chemistry and Biochemistry, Medical University of Sofia, Sofia, 2 Zdrave Str., 1431 Sofia, Bulgaria

74 Department of Oncology, Cross Cancer Institute, University of Alberta, 11560 University Avenue, Edmonton, Alberta, Canada T6G 1Z2

75 Division of Radiation Oncology, Cross Cancer Institute, 11560 University Avenue, Edmonton, Alberta, Canada T6G 1Z2

76 Molecular Endocrinology Laboratory, Department of Cellular and Molecular Medicine, KU Leuven, BE-3000, Belgium

77 Division of Cancer Sciences, Manchester Cancer Research Centre, Faculty of Biology, Medicine and Health, Manchester Academic Health Science Centre, NIHR Manchester Biomedical Research Centre, Health Innovation Manchester, Univeristy of Manchester, M13 9WL

78 The University of Surrey, Guildford, Surrey, GU2 7XH, UK

79 Genetic Oncology Unit, CHUVI Hospital, Complexo Hospitalario Universitario de Vigo, Instituto de Investigación Biomédica Galicia Sur (IISGS), 36204, Vigo (Pontevedra), Spain

80 Department of Urology, Erasmus University Medical Center, 3015 CE Rotterdam, The Netherlands

81 "Exposome and Heredity", CESP (UMR 1018), Faculté de Médecine, Université Paris-Saclay, Inserm, Gustave Roussy, Villejuif

82 Clinical Gerontology Unit, University of Cambridge, Cambridge, CB2 2QQ, UK

83 Division of Epidemiology, Department of Internal Medicine, University of Utah School of Medicine, Salt Lake City, Utah 84132, USA

84 George E. Wahlen Department of Veterans Affairs Medical Center, Salt Lake City, Utah 84148, USA

85 Department of Laboratory Medicine and Pathology, Mayo Clinic, Rochester, MN 55905, USA

86 Nuffield Department of Population Health, University of Oxford, United Kingdom

87 Program in Genetic Epidemiology and Statistical Genetics, Department of Epidemiology, Harvard School of Public Health, Boston, MA, USA

88 Division of Epidemiology, Department of Medicine, Vanderbilt University Medical Center, 2525 West End Avenue, Suite 800, Nashville, TN 37232 USA

89 International Epidemiology Institute, Rockville, MD 20850, USA

90 Department of Epidemiology and Biostatistics, School of Public Health, Imperial College London, SW7 2AZ, UK
